# Predicting Postoperative Delirium in Older Patients: a multicenter retrospective cohort study

**DOI:** 10.1101/2024.03.13.24303920

**Authors:** Shun-Chin Jim Wu, Nitin Sharma, Anne Bauch, Hao-Chun Yang, Jasmine L. Hect, Christine Thomas, Sören Wagner, Bernd R. Förstner, Christine A.F. von Arnim, Tobias Kaufmann, Gerhard W. Eschweiler, Thomas Wolfers, the PAWEL Study Group

**Affiliations:** Department of Psychiatry and Psychotherapy, University of Tübingen, Germany; German Centre for Mental Health, Tübingen, Germany; Department of Psychiatry, University of Cambridge, UK; Department of Neurological Surgery, University of Pittsburgh School of Medicine, Pittsburgh Pennsylvania, USA; Department of Geriatric Psychiatry and Psychotherapy, Klinikum Stuttgart, Stuttgart, Germany; Department of Anesthesiology, Klinikum Stuttgart, Stuttgart, Germany; Department of Anesthesiology and Intensive Care, School of Medicine and Health, Klinikum Rechts der Isar, Technical University of Munich, Munich, Germany; Social and Preventive Medicine, Department of Sports and Health Sciences, University of Potsdam, Potsdam, Germany; Department of Geriatrics, University Medical Center Göttingen, Göttingen, Germany; German Center for Cardiovascular Research (DZHK), Göttingen, Germany; Geriatric Center, Universitätsklinikum Tübingen, Tübingen, Germany; Norwegian Centre for Mental Disorders Research, University of Oslo, Norway; Donders Institute, Radboud University Nijmegen, The Netherlands

**Keywords:** Postoperative delirium, Machine learning, Explainability, Preoperative cognition, Geriatrics

## Abstract

**Background:** The number of elective surgeries for older individuals is on the rise globally. Machine learning may improve risk assessment with an impact on surgical planning and postoperative care. Preoperative cognitive assessment may facilitate early identification of postoperative delirium (POD). This study aims to estimate the predictive ability of machine learning models for POD using pre-and/or perioperative features, with a specific focus on adding neuropsychological assessments prior to surgery.

**Materials and Methods:** This retrospective cohort study analyzed data from the multicenter PAWEL study and its PAWEL-R substudy, encompassing older patients (≥70 years) undergoing elective surgeries across five medical centers from July 2017 to April 2019. A total of 1624 patients were included, with POD diagnosis made before discharge. Data included demographics, clinical, surgical, and neuropsychological features collected pre- and perioperatively. Machine learning model performance was evaluated using the area under the receiver operating characteristic curve (AUC), with permutation testing for significance and SHapley Additive exPlanations (SHAP) to identify effective neuropsychological assessments.

**Results:** In this cohort of 1624 patients, 52.3% (N=850) were male, with a mean [SD] age of 77.9 [4.9] years. Predicting POD before surgery using demographic, clinical, surgical, and neuropsychological features achieved an AUC of 0.79. Incorporating all pre- and perioperative features into the model yielded a slightly higher AUC of 0.82, with no significant difference observed (*P*= .19). Notably, cognitive factors alone were not strong predictors (AUC=0.61). However, specific tests within neuropsychological assessments, such as the Montreal Cognitive Assessment memory subdomain and Trail Making Test Part B, were found to be crucial for prediction according to SHAP analysis.

**Conclusion and Relevance:** Preoperative risk prediction for POD can increase risk awareness in presurgical assessment and improve postoperative management in patients with a high risk for delirium.

**Highlights:**

1. Analyzed 1624 older patients (≥70 years) undergoing elective surgeries across five medical centers from July 2017 to April 2019.
2. Established machine learning model to predict postoperative delirium before surgery.
3. Preoperative cognition enhances predictive performance, comparable to models incorporating all pre- and perioperative features.
4. Montreal Cognitive Assessment memory subdomain and Trail Making Test Part B drive the cognition-based prediction.
5. Perioperative surgical features, such as the duration of the surgery, are important predictors.

## Introduction

In an aging society, there is a rising demand for elective surgeries due to the changing healthcare needs of older people ^1–3^. However, this increase in elective surgeries raises concerns about additional adverse outcomes, particularly given the unique challenges posed by aging, such as pre-existing health conditions, disease sequelae, and diminished physiological reserves ^4,5^. Meeting the growing demand for elective surgeries among the elderly requires a comprehensive strategy, including thorough preoperative assessments, personalized care plans, and continuous post-operative support ^6–8^. In this study, we systematically assess the potential benefits of utilizing machine learning techniques to predict the risk of postoperative delirium (POD) based on a diverse range of features.

POD, characterized by acute and fluctuating inattention with alterations in thinking or consciousness after surgery, affects 12% to 51% of older patients, with incidence varying by surgical procedures and regions ^9^. POD in older patients is linked to heightened rehospitalization rates, persistent postoperative cognitive dysfunction, increased incidence of dementia, and elevated mortality ^10–12^. Some studies have shown significant associations between POD and factors collected before or during surgery (pre- or perioperatively) ^13,14^. These factors could aid in predicting the POD ^15,16^. However, different pre- and perioperative feature categories may vary in their importance for POD risk assessment. Demographic factors, such as age and sex, are critical for assessing POD risk ^17–23^. Clinical data, including blood samples and chronic disease medication, are also predictive ^24–26^. The Type and length of surgery and anesthesia are indispensable for POD prediction ^18,27^. Although preoperative neuropsychological assessments, like the Mini-Mental State Examination (MMSE) and others ^28–30^, help identify at-risk patients for early risk mitigation ^31–33^, these evaluations are not yet incorporated in clinical routine despite experts’ recommendations ^34^. The early identification of POD risk factors enables clinicians to proactively mitigate the occurrence of POD ^35^ and might affect patient’s decision before non-emergent surgery. Therefore, identifying preoperative risk factors for POD is crucial in facilitating personalized surgical risk assessment before surgery ^36,37^. Moreover, stratifying POD risks remains challenging due to its multifactorial origins ^18,22,38^. The precise categories of preoperative and perioperative features with superior predictive capabilities for POD remain unknown and unvalidated ^39^.

In this study we (1) predict POD by employing a machine learning approach utilizing diverse pre- and perioperative features from a large multicenter cohort of older patients and (2) conduct a comparative analysis of independent models using various data categories, including demographic, clinical, surgical, and neuropsychological features. This study provides a comprehensive assessment of predictors for POD thereby enhancing presurgical awareness of risk and postoperative management in older patients.

## Materials and Methods

### Participants

The study utilized the cohort from the PAWEL study (Patientensicherheit, Wirtschaftlichkeit und Lebensqualität bei elektiven Operationen, English: Patient safety, Efficiency and Life quality in elective surgery) and its PAWEL-R sub-study (R for risk) to predict POD using machine learning models, departing from original statistical methods in the previous studies. Patients were recruited from five major medical institutions in Germany (three university hospitals: Tübingen, Freiburg, and Ulm, and two tertiary medical centers: Stuttgart and Karlsruhe) between July 31, 2017, and April 12, 2019 ^13,40,41^. The study adheres to the guidelines outlined in the STROCSS criteria ^42^.

Participants included patients aged 70 and older undergoing elective surgery (joint, spine, vessels, heart, lung, abdomen, urogenital system, and other organs) with an expected surgical duration exceeding 60 minutes. Exclusion criteria covered patients unable to communicate effectively in German, those undergoing emergency surgery, severe dementia with MMSE < 15 or Montreal Cognitive Assessment (MOCA) < 8, or an estimated survival time less than 15 months. A stepped-wedge cluster randomized design was employed for equitable intervention group allocation ^40,41^. The study, initially including 1631 patients, conducted thorough postoperative assessments within one week after surgery but before discharge. To ensure accuracy in predicting POD diagnosis, patients without POD diagnosis before discharge (N=7) were excluded, resulting in 1624 patients included in prediction models. The study reported 23.1% of patients diagnosed with POD, with detailed group comparisons available in eTables 1 and 2.

### Measures

The PAWEL and PAWEL-R studies extensively evaluated elective surgery patients to identify risk factors and outcomes related to POD. Diagnosis involved the Confusion Assessment Method (I-CAM) algorithm and chart review within the first postoperative week or until discharge. Demographic data, including age, sex, education, alcohol/smoking habits, living arrangement, and hospital location, were collected preoperatively. Neuropsychological assessments such as MOCA, Trial Making Test (TMT) parts A and B, digit span backwards, Subjective Memory Impairment (SMI), and Patient Health Questionnaire-4 (PHQ-4) were conducted at admission. Clinical profiles included blood samples, past medical histories including pre-existing dementia and previous delirium history, and baseline assessments. Surgical information covered types of surgery, anesthesia, and perioperative events. Preoperative and perioperative features were analyzed for predictive capacity at two time points. Models were developed and tested with both features and preoperative features alone. Additionally, models were compared with and without neuropsychological assessments to assess their contribution to predictive performance. The study aimed to identify effective feature combinations for predicting POD diagnosis by comparing predictive performance across different feature combinations from the two time points.

### Preprocessing, imputation, and features

Features with more than 20% missing data, such as albumin level (60.28%) and depth of anesthesia (75.37%), were excluded ^16^. Information about missing values is available in eTable 3. Data imputation involved using the mean value for continuous variables and random sampling from the original probability distribution for discrete and binary variables within cross-validation folds. Blood samples, including hemoglobin, sodium levels, and C-reactive protein (CRP), were interpreted following clinical guidelines in line with standard practice ^43^. For instance, hemoglobin levels of less than 12 g/dL were considered indicative of anemia, while sodium levels of less than 135 mmol/L and more than 145 mmol/L were associated with hyponatremia and hypernatremia, respectively. Similarly, CRP levels greater than 3 mg/L indicated an increased CRP level, while values within the normal range were not considered clinically significant. Redundant features with a perfect correlation were excluded. Non-binary categorical features, including location, SMI, types of anesthesia, and surgery, were one-hot encoded because they had no natural ordinal relationship among their categories, and assigning numerical labels to them could introduce bias or incorrect assumptions in the model.

Seventy features were used, divided into preoperative (51) and perioperative (19) categories. Preoperative features encompassed demographic (7), clinical (23), surgical categories (6), and neuropsychological assessments (15). Perioperative features included clinical (4) and surgical (15) categories. Different combinations of features were compared, including preoperative only, preoperative and perioperative, and sub-feature sets of preoperative features. The study evaluated model performance, feature category effectiveness, and the additional benefits of preoperative neuropsychological assessments.

The study aimed to develop a prediction model in a naturalistic setting. Information regarding interventions was included only as a sensitivity analysis to demonstrate its potential impact on the prediction model. The potential imbalance of dataset for all models was tested by oversampling with the Synthetic Minority Oversampling Technique (SMOTE) ^44^. Possible entry errors in clinical assessments were identified by mean imputation of data points exceeding five times the standard deviation on a respective measure. Model performances were estimated and compared to the original predictions, which included those datapoints. Various sensitivity analyses as described in the supplement were performed.

### Machine learning models, performance evaluation and feature importance

Machine Learning models were used to predict POD, including logistic regression, support vector machines, random forest, and gradient boosting without hyperparameters tuning using the scikit-learn library version 1.2.2 ^45^ and the Xgboost library version1.7.3 ^46^. Independent variables were feature variables, while the dependent variable was POD diagnosis, as illustrated in Figure 1-B. Five-fold cross-validation, with balanced labels across folds, measured model performance at testing using the area under the receiver operating characteristic curve (AUC) as the primary metric. Additional metrics included precision, recall, sensitivity, specificity, balanced accuracy, and area under the precision-recall curve presented in eTable 4. Permutation testing assessed AUC values compared to random chance. POD diagnosis labels were shuffled for random chance, and AUC values were measured 1000 times. The p-value compared the original AUC value to the distribution of permuted AUC values. The difference between models was assessed similarly, calculating the difference between the AUC values of two models with permuted labels^47^. The SHapley Additive exPlanations (SHAP) values were used to assess feature importance. SHAP measures each feature’s influence on the model’s prediction. Positive SHAP values increased the probability of POD, while negative values decreased it. All features’ average absolute SHAP value indicated contributions to the model’s prediction. The SHAP library version 0.41.0 in Python was employed ^48^. All preprocessing steps and analyses are available on GitHub upon publication from Sharma forked to https://github.com/MHM-lab. The data can be requested by addressing the PAWEL consortium.

**Figure 1.**
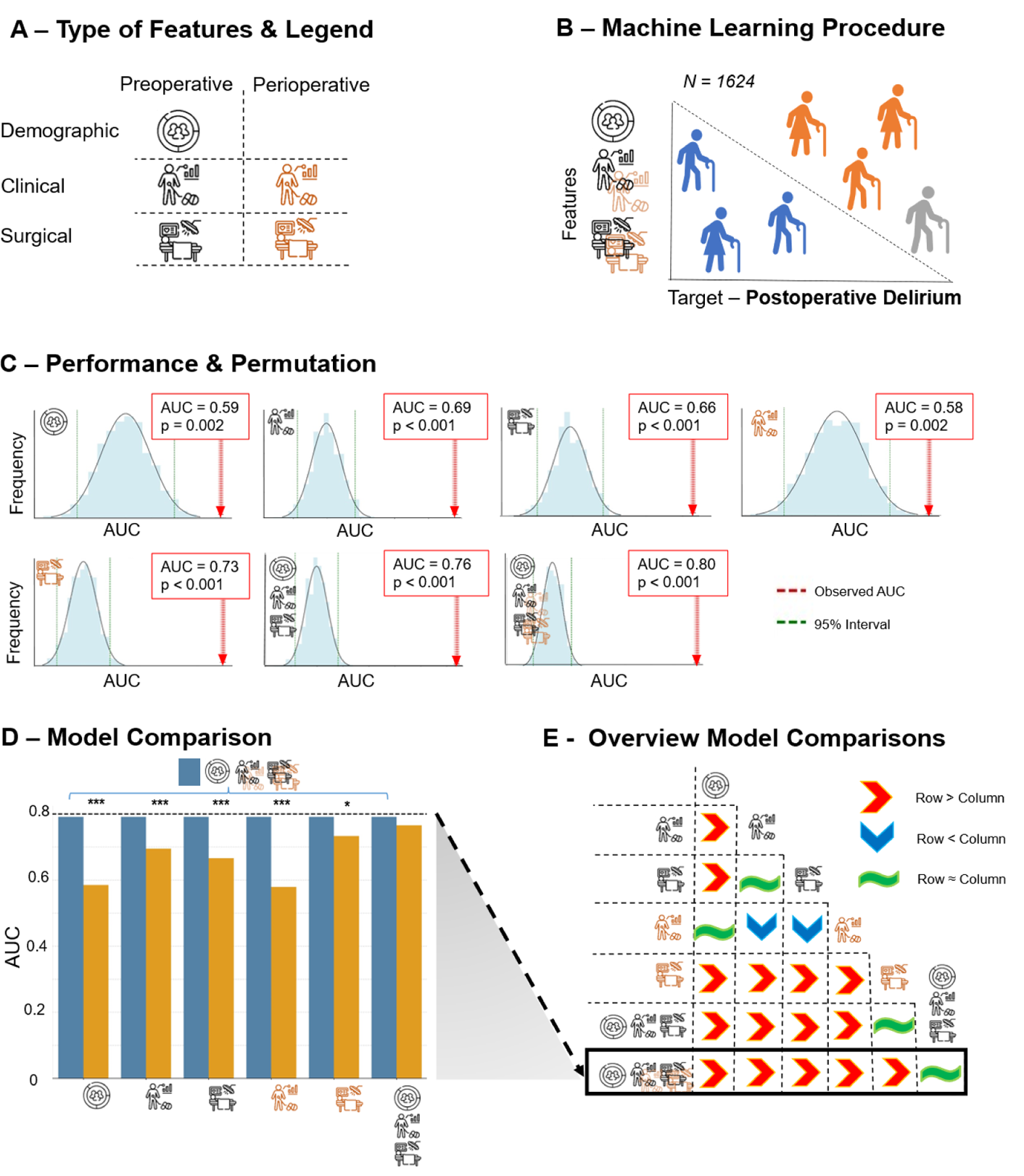
Individual feature categories perform worse than combined feature categories pre- and perioperatively and POD can effectively be predicted prior to surgery (see back box). (A) The symbols are consistently used in this study and represent the different types of features. (B) We employed a random forest classifier with 5-fold cross-validation to forecast postoperative delirium (POD) onset in 1624 older individuals. Among them, 23.1% experienced POD. Orange indicates patients with POD, blue represents those without POD, and grey signifies patients predicted to have POD by the algorithm. (C) Through permutation testing we demonstrated that both individual and combined features have predictive power for POD beyond chance. (D) No significant difference in their performance was found when comparing models utilizing preoperative features alone and those incorporating both preoperative and perioperative features (blue bar). The blue bar represents the reference model, while the yellow bars represent the comparison models. The asterisks (*) denote statistical significance levels for the difference in the Area Under the receiver operating characteristic Curve (AUC) between pre-and perioperative features and other feature types, calculated over 1000 permutations: \**P* <.05, \*\**P* ≤.01, \*\*\**P* ≤.001. (E) A comparative analysis of different feature combinations based on their respective AUC values was conducted. Combined feature sets consistently outperformed individual feature types. Among individual feature types, surgical data from the day of the operation emerged as the strongest predictor category for POD.

## Results

### Predicting POD with combined, pre-, and perioperative features

Evaluating four classifiers, comparable performances were observed (eFigure 1). Here, we accentuate the findings obtained from random forest due to its marginally better performance across most models. The models incorporating combined and independent pre- and perioperative features exhibited robust performance, as evidenced by AUC values surpassing chance levels (Figure 1-C and eTables 4 and 5) and eFigure 2 displays the performance of these models through receiver operating characteristic curves. Notably, the model using only preoperative features (Pre-Op) achieved an AUC of 0.76, comparable to a model incorporating both pre- and perioperative features (Pre and Peri-Op), which had an AUC of 0.80, showing no significant difference (Figure 1-D). The independent model exclusively utilizing perioperative surgical features demonstrated an AUC value of 0.73 (Figure 1-E). It is noteworthy that longer cut-to-suture time (surgical duration) and increased equipment usage duration were associated with a higher likelihood of POD, as shown in eFigure 5. Further details, including pairwise comparisons and p-values for differences in AUC values, can be found in eFigure 3 and eTable 6.

**Figure 2.**
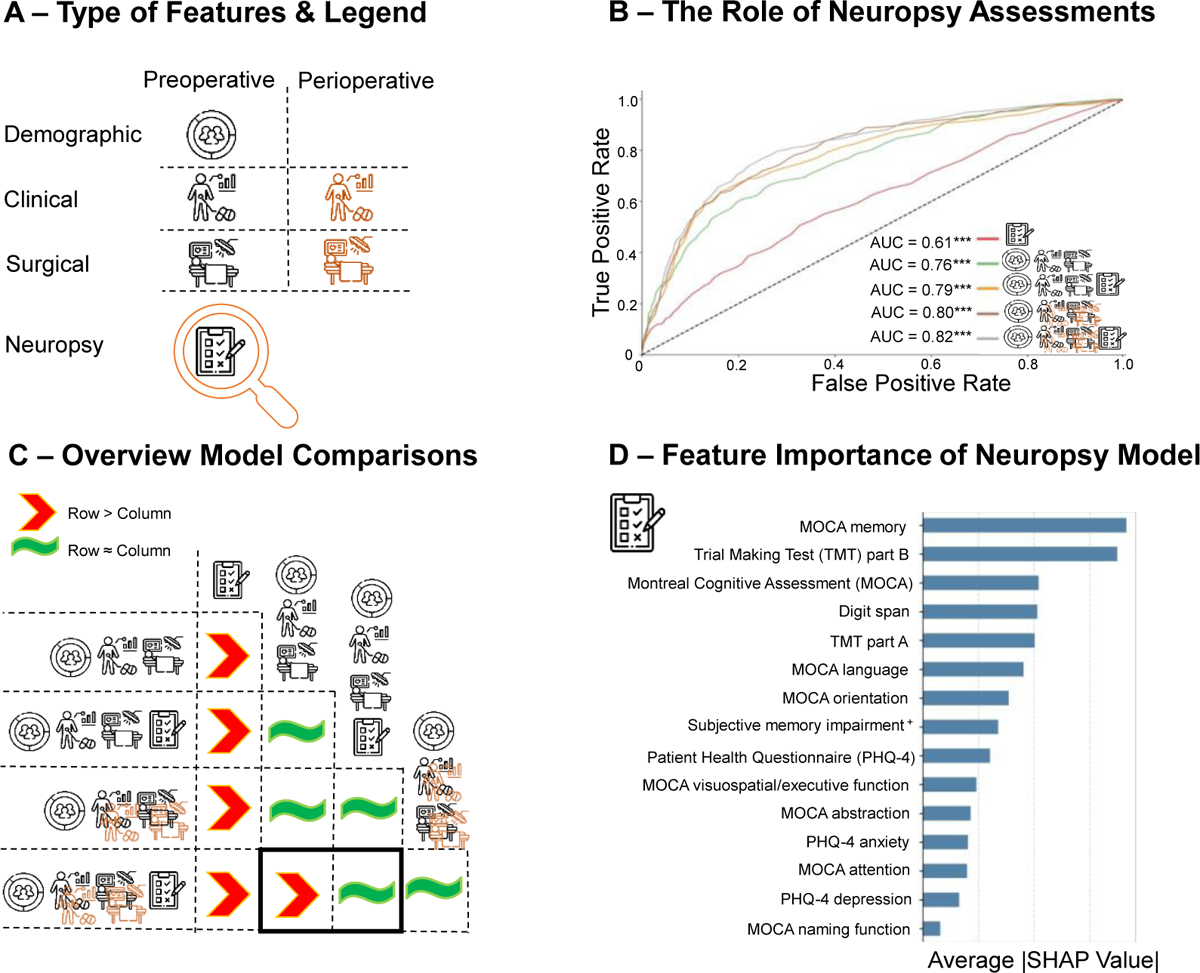
Cognition, in isolation, demonstrates limited predictive power. However, when combined with preoperative features the performance matches that of all pre- and perioperative features (see back box). (A) The predictive capability of preoperative neuropsychological tests was assessed for postoperative delirium (POD) individually and in combination with other features. (B) Our analysis, as depicted in the receiver operating characteristic curve, demonstrated the successful prediction of POD by preoperative neuropsychological tests. However, when combined with pre- and perioperative data, the performance gains were only marginal. Asterisks (*) denote the Area Under the receiver operating characteristic Curve (AUC) values, which were assessed against chance permutation testing, revealing a significance (\*\*\**P* ≤.001). (C) The AUC values of neuropsychological tests were compared to those integrated into the pre- and perioperative features. While preoperative neuropsychological tests did not prove helpful in predicting POD after surgery, they contributed to POD prediction before the surgical procedure. (D) SHapley Additive exPlanations (SHAP) was employed to assess feature importance. The feature importance was based on the average absolute value of the SHAP score within the preoperative features. Specific neuropsychological tests, such as the Montreal Cognitive Assessment (MOCA) memory subdomain and Trail Making Tests (TMT) part B, were particularly influential.

### Addition of preoperative neuropsychological assessments

The model using preoperative neuropsychological assessments exclusively, exhibited AUC values of 0.61. Integrating neuropsychological assessments into the model, utilizing both pre- and perioperative features (Pre and Peri-Op + NeuroPsy), led to a slight improvement in the AUC, reaching 0.82. This enhancement elevated the model to the top-performing one (Figure 2-B). Adding neuropsychological assessments to the preoperative model (Pre-Op + NeuroPsy) improved the AUC from 0.76 to 0.79, which resulted in a non-significant model comparison including a model based on all pre- and perioperative features (Pre and Peri-Op) with the AUC of 0.80 (Figure 2-C). Specifically, poorer performance in the MOCA memory subdomain and TMT part B before surgery indicated a higher likelihood of POD, as illustrated in Figure 2-D and eFigure 6. For detailed pairwise comparisons of AUC values and corresponding p-values, please refer to eFigure 4 and eTable 6. Including intervention allocation information had no discernible impact on predictive performance (eFigure 7 and eTables 4 and 7). Lastly, there was no difference in AUC between models with and without oversampling (eFigure 8) and outliers or erroneous data didn’t affect our models’ performance (eFigure 9).

## Discussion

Leveraging machine learning, we forecasted the occurrence of POD after elective surgeries through a combination of preoperative and perioperative features with a large multicenter cohort. Although our models showcased robust preoperative performance, it was the perioperative surgical data that proved to be the most potent sole predictors category of POD. The integration of preoperative neuropsychological tests yielded an enhancement in the AUC comparable to the model with all pre- and perioperative features, notably influenced by the MOCA memory subdomain and TMT part B, as elucidated by model explanations. This integrated analysis improves conventional clinical risk profiling, furnishing superior predictive capacity with promising implications for surgical planning in the era of machine learning-assisted healthcare and empowers the prioritization of pivotal features in future work.

### Perioperative surgical features

Perioperative surgical features, including those on the day of the operation, emerged as the best solo predictors category for POD, as illustrated in Figure 1-E. Their significant SHAP values highlighted vital factors such as ventilation duration, surgical procedures overall duration, and central venous catheter use, anticoagulants, oxygen use duration, as depicted in eFigure 5-B. Prolonged individual surgical durations with longer equipment use were associated with a higher risk of POD. This association was particularly notable in surgeries involving the use of a heart-lung machine (eFigure 5-A), with significantly more patients experiencing POD undergoing cardiopulmonary bypass (47.1%) compared to those not experiencing POD (17.8%) (eTable 1). Consistent with our findings, even a 30-minute increase in surgery duration corresponded to a 6% rise in POD risk ^27^, and this risk is further elevated in prolonged use of cardiopulmonary bypass during cardiac surgeries ^49^. This might be explained by potential hypoperfusion or micro-embolism in cardiac surgeries, elevating the risk of POD ^50^.

In contrast to the significance of the surgery type during the preoperative phase, our analysis revealed that the duration of individual surgery with ventilation emerged as a more influential factor in predicting POD. Overall, our findings highlight the critical importance of cardiovascular risk measures and surgical metrics in POD prediction. Looking ahead, the integration of real-time predictive technology into surgical workflows holds promise. This advancement could potentially facilitate on-the-fly predictions during surgery, enabling timely adjustments to medication or non-pharmacological intervention to mitigate potential adverse outcomes associated with surgical interventions. Our study emphasizes the substantial value of information gleaned from measures taken during surgery, shedding light on their crucial role in enhancing our understanding and prediction of POD.

### Enhanced POD prediction prior to surgery through neuropsychological assessments

The preoperative model for predicting POD demonstrated effectiveness, which is in line with previous work ^16,36,37^. Although incorporating perioperative data slightly improved POD predictions, it’s crucial to note that this information is only available during and immediately after surgery. This limitation prevents surgical planning and decision-making beforehand, allowing only for adjustments in real-time. Therefore, augmenting the predictive performance of algorithms by incorporating data that can be gathered prior to a surgical procedure is important, as it allows for integrated surgical planning and informed decision making prior to any invasive or surgical procedure. In our study, preoperative neuropsychological assessments were predictive above chance, however, the added benefit to combined pre- and perioperative features was modest (Figure 2-B). However, the absence of preoperative neuropsychological tests reduced the performance of POD prediction in the context of models solely relying on preoperative features (Figure 2-C), which is critical given that surgical and postoperative management could be optimized. Preoperative models with neuropsychological tests effectively predicted POD before surgery substantiating previous observations ^13,28,51^. This could be attributed to preoperative neuropsychological tests revealing subtle cognitive deficits, despite less than 2% of dementia diagnoses before surgery in this study cohort (eTable 1). These deficits may progress into POD. In line with this explanation, timely preoperative cognitive interventions can mitigate the risk of POD and long-term cognitive dysfunction after cardiac surgeries^52,53^. Additionally, patients with pre-existing cognitive decline face increased risks of other postoperative complications ^54^. Consequently, baseline neuropsychological assessments are valuable for improving the prediction of POD although and as shown in this study with modest additional effects.

Selecting suitable neuropsychological assessments for clinical use is crucial. Our study identified the MOCA memory subdomain and TMT part B as effective indicated by their average absolute SHAP values (Figure 2-D). Low scores on the MOCA memory subdomain and longer test times on the TMT Part B indicate poor cognitive performance and executive dysfunction (eFigure 6). These tests are crucial for predicting POD risk, as demonstrated in a prior prediction study ^15^. Patients with mild cognitive impairment at baseline are more likely to develop POD ^25^, while good preoperative cognitive performance is protective against POD ^18^. Previous studies have often used the MMSE ^28,31,55^, Clock-Drawing Test ^20^, or MOCA score as preoperative risk factors ^56^. Critically, we replicated the strong association between baseline MOCA and POD risk in the previous theory-driven PAWEL-R study with a larger cohort^13^. These findings offer a thorough understanding of the efficacy of individual preoperative neuropsychological tests in predicting POD. By conducting comprehensive assessments of pre-existing risks, we may unlock new avenues for optimizing surgical planning and postoperative management. Our study underscores the added albeit moderate advantage of evaluating cognition, emphasizing its importance, and advocating for its inclusion in future developments aimed at refining preoperative risk assessments.

### Limitations and recommendation

The study exhibits several limitations that require consideration. First, interpreting SHAP values warrants caution ^57^, as is generally the case for methods using model explanations in medicine ^58,59^. Unstable explanations are not uncommon for complex models trained on large datasets ^60,61^. While the ranking of importance may fluctuate, features with higher mean absolute SHAP values generally maintain consistent attributions. Second, we harnessed cognitive data derived from standardized assessments. However, it’s crucial to note that these procedures, while standardized, often remain non-digitalized. This presents significant untapped potential for future advancements in the realm of risk evaluation before surgical procedures. Third, while our study included a wide range of preoperative neuropsychological assessments, it’s not an exhaustive list. As neuropsychological assessments are time-consuming and require training for assessors to conduct accurate tests and interpret results. Therefore, incorporating semi-automatic assessments of cognition ^62^ may prove advantageous and a relevant direction for future research.

## Conclusion and relevance

We showcase robust predictions of POD in older patients, relying on a combination of pre- and perioperative features. This study further underscores the feasibility of predicting POD prior to surgery and the additional incorporation of preoperative neuropsychological assessments. Our research contributes to a heightened comprehension of predictors for POD prioritizing distinct variable dimensions, thereby guiding a more targeted approach in addressing POD risk predictions in clinical practice.

## Data Availability

The data can be requested by addressing the PAWEL consortium.

## Acknowledgements

Assistance with the study: We would like to thank the PAWEL and PAWEL-R Study group and the patients’ participation in this study, making the work possible. Financial support and sponsorship: This work was supported by grant VF1_2016-201 from the Innovationsfonds (fund of the Gemeinsamer Bundesausschuss, GBA) as well as the German Research Foundation (DFG) Emmy Noether with reference 513851350 (TW) and the Cluster of Excellence with reference 390727645 (TW). This work was supported by the BMBF-funded de.NBI Cloud within the German Network for Bioinformatics Infrastructure (de.NBI) (031A532B, 031A533A, 031A533B, 031A534A, 031A535A, 031A537A, 031A537B, 031A537C, 031A537D, 031A538A). Conflicts of interest: CT received honoraria from serving on the scientific advisory board of Roche, a research grant from the gemeinsame Bundesausschuß der Krankenkassen and has received funding for travel and speaker honoraria from several hospitals for scientific education. GWE has nothing to disclosure. CAFvA received honoraria from serving on the scientific advisory board of Biogen, Roche, Novo Nordisk, Biontech, MindAhead UG and Dr. Willmar Schwabe GmbH &Co. KG and has received funding for travel and speaker honoraria from Lilly, Biogen, Roche diagnostics AG, Novartis, Medical Tribune Verlagsgesellschaft mbH, Landesvereinigung für Gesundheit und Akademie für Sozialmedizin Niedersachsen e. V., FomF GmbH | Forum für medizinische Fortbildung and Dr. Willmar Schwabe GmbH &Co. KG and has received research support from Roche diagnostics AG. Presentation: none.

## References

1. Deng C, Mitchell S, Paine SJ, Kerse N. Retrospective analysis of the 13-year trend in acute and elective surgery for patients aged 60 years and over at Auckland City Hospital, New Zealand. J Epidemiol Community Health. Jan 2020;74(1):42–47. doi:10.1136/jech-2019-212283

2. Lee YZ, Dharmawan A, Zhang X, Chua DYC, Low JK. The changing landscape of general surgery in the elderly – trends over a decade in a tertiary centre in Singapore. ANZ Journal of Surgery. 2022;92(9):2018–2024. 10.1111/ans.17674

3. Lutz W, Sanderson W, Scherbov S. The coming acceleration of global population ageing. Nature. 2008/02/01 2008;451(7179):716–719. doi:10.1038/nature06516

4. Glance LG, Benesch CG, Holloway RG, et al. Association of Time Elapsed Since Ischemic Stroke With Risk of Recurrent Stroke in Older Patients Undergoing Elective Nonneurologic, Noncardiac Surgery. JAMA Surgery. 2022;157(8):e222236–e222236. doi:10.1001/jamasurg.2022.2236

5. Graham LA, Hawn MT. Managing Competing Risks for Surgical Patients With Complex Medical Problems—Considering Confounding. JAMA Surgery. 2024;159(2):149–150. doi:10.1001/jamasurg.2023.5952

6. Partridge JSL, Harari D, Martin FC, Dhesi JK. The impact of pre-operative comprehensive geriatric assessment on postoperative outcomes in older patients undergoing scheduled surgery: a systematic review. Anaesthesia. 2014;69(s1):8–16. 10.1111/anae.12494

7. Kim KI, Park KH, Koo KH, Han HS, Kim CH. Comprehensive geriatric assessment can predict postoperative morbidity and mortality in elderly patients undergoing elective surgery. Arch Gerontol Geriatr. May-Jun 2013;56(3):507–12. doi:10.1016/j.archger.2012.09.002

8. Aceto P, Antonelli Incalzi R, Bettelli G, et al. Perioperative Management of Elderly patients (PriME): recommendations from an Italian intersociety consensus. Aging Clin Exp Res. Sep 2020;32(9):1647–1673. doi:10.1007/s40520-020-01624-x

9. Whitlock EL, Vannucci A, Avidan MS. Postoperative delirium. Minerva Anestesiol. Apr 2011;77(4):448–56.

10. Witlox J, Eurelings LS, de Jonghe JF, Kalisvaart KJ, Eikelenboom P, van Gool WA. Delirium in elderly patients and the risk of postdischarge mortality, institutionalization, and dementia: a meta-analysis. Jama. Jul 28 2010;304(4):443–51. doi:10.1001/jama.2010.1013

11. Vasilevskis EE, Han JH, Hughes CG, Ely EW. Epidemiology and risk factors for delirium across hospital settings. Best Pract Res Clin Anaesthesiol. Sep 2012;26(3):277–87. doi:10.1016/j.bpa.2012.07.003

12. Gleason LJ, Schmitt EM, Kosar CM, et al. Effect of Delirium and Other Major Complications on Outcomes After Elective Surgery in Older Adults. JAMA Surgery. 2015;150(12):1134–1140. doi:10.1001/jamasurg.2015.2606

13. Eschweiler GW, Czornik M, Herrmann ML, et al. Presurgical Screening Improves Risk Prediction for Delirium in Elective Surgery of Older Patients: The PAWEL RISK Study. Front Aging Neurosci. 2021;13:679933. doi:10.3389/fnagi.2021.679933

14. Susano MJ, Scheetz SD, Grasfield RH, et al. Retrospective Analysis of Perioperative Variables Associated With Postoperative Delirium and Other Adverse Outcomes in Older Patients After Spine Surgery. J Neurosurg Anesthesiol. Oct 2019;31(4):385–391. doi:10.1097/ana.0000000000000566

15. Sadlonova M, Hansen N, Esselmann H, et al. Preoperative Delirium Risk Screening in Patients Undergoing a Cardiac Surgery: Results from the Prospective Observational FINDERI Study. Am J Geriatr Psychiatry. Dec 29 2023;doi:10.1016/j.jagp.2023.12.017

16. Xue B, Li D, Lu C, et al. Use of Machine Learning to Develop and Evaluate Models Using Preoperative and Intraoperative Data to Identify Risks of Postoperative Complications. JAMA Netw Open. Mar 1 2021;4(3):e212240. doi:10.1001/jamanetworkopen.2021.2240

17. Chaiwat O, Chanidnuan M, Pancharoen W, et al. Postoperative delirium in critically ill surgical patients: incidence, risk factors, and predictive scores. BMC Anesthesiol. Mar 20 2019;19(1):39. doi:10.1186/s12871-019-0694-x

18. Greaves D, Psaltis PJ, Davis DHJ, et al. Risk Factors for Delirium and Cognitive Decline Following Coronary Artery Bypass Grafting Surgery: A Systematic Review and Meta-Analysis. J Am Heart Assoc. Nov 17 2020;9(22):e017275. doi:10.1161/jaha.120.017275

19. Jankowski CJ, Trenerry MR, Cook DJ, et al. Cognitive and functional predictors and sequelae of postoperative delirium in elderly patients undergoing elective joint arthroplasty. Anesth Analg. May 2011;112(5):1186–93. doi:10.1213/ANE.0b013e318211501b

20. Vasilian CC, Tamasan SC, Lungeanu D, Poenaru DV. Clock-Drawing Test as a Bedside Assessment of Post-operative Delirium Risk in Elderly Patients with Accidental Hip Fracture. World J Surg. May 2018;42(5):1340–1345. doi:10.1007/s00268-017-4294-y

21. Wang CG, Qin YF, Wan X, Song LC, Li ZJ, Li H. Incidence and risk factors of postoperative delirium in the elderly patients with hip fracture. J Orthop Surg Res. Jul 27 2018;13(1):186. doi:10.1186/s13018-018-0897-8

22. Wu J, Yin Y, Jin M, Li B. The risk factors for postoperative delirium in adult patients after hip fracture surgery: a systematic review and meta-analysis. Int J Geriatr Psychiatry. Jan 2021;36(1):3–14. doi:10.1002/gps.5408

23. Wang H, Guo X, Zhu X, et al. Gender Differences and Postoperative Delirium in Adult Patients Undergoing Cardiac Valve Surgery. Front Cardiovasc Med. 2021;8:751421. doi:10.3389/fcvm.2021.751421

24. Ayob F, Lam E, Ho G, Chung F, El-Beheiry H, Wong J. Pre-operative biomarkers and imaging tests as predictors of post-operative delirium in non-cardiac surgical patients: a systematic review. BMC Anesthesiol. Feb 23 2019;19(1):25. doi:10.1186/s12871-019-0693-y

25. Kazmierski J, Banys A, Latek J, et al. Mild cognitive impairment with associated inflammatory and cortisol alterations as independent risk factor for postoperative delirium. Dement Geriatr Cogn Disord. 2014;38(1-2):65–78. doi:10.1159/000357454

26. Kassie GM, Nguyen TA, Kalisch Ellett LM, Pratt NL, Roughead EE. Do Risk Prediction Models for Postoperative Delirium Consider Patients’ Preoperative Medication Use? Drugs Aging. Mar 2018;35(3):213–222. doi:10.1007/s40266-018-0526-6

27. Ravi B, Pincus D, Choi S, Jenkinson R, Wasserstein DN, Redelmeier DA. Association of Duration of Surgery With Postoperative Delirium Among Patients Receiving Hip Fracture Repair. JAMA Netw Open. Feb 1 2019;2(2):e190111. doi:10.1001/jamanetworkopen.2019.0111

28. Liu J, Li J, He J, Zhang H, Liu M, Rong J. The Age-adjusted Charlson Comorbidity Index predicts post-operative delirium in the elderly following thoracic and abdominal surgery: A prospective observational cohort study. Front Aging Neurosci. 2022;14:979119. doi:10.3389/fnagi.2022.979119

29. Lin X, Liu F, Wang B, et al. Subjective Cognitive Decline May Be Associated With Post-operative Delirium in Patients Undergoing Total Hip Replacement: The PNDABLE Study. Front Aging Neurosci. 2021;13:680672. doi:10.3389/fnagi.2021.680672

30. Segernäs A, Skoog J, Ahlgren Andersson E, Almerud Österberg S, Thulesius H, Zachrisson H. Prediction of Postoperative Delirium After Cardiac Surgery with A Quick Test of Cognitive Speed, Mini-Mental State Examination and Hospital Anxiety and Depression Scale. Clin Interv Aging. 2022;17:359–368. doi:10.2147/cia.S350195

31. Veliz-Reissmüller G, Agüero Torres H, van der Linden J, Lindblom D, Eriksdotter Jönhagen M. Pre-operative mild cognitive dysfunction predicts risk for post-operative delirium after elective cardiac surgery. Aging Clin Exp Res. Jun 2007;19(3):172–7. doi:10.1007/bf03324686

32. Cao SJ, Chen D, Yang L, Zhu T. Effects of an abnormal mini-mental state examination score on postoperative outcomes in geriatric surgical patients: a meta-analysis. BMC Anesthesiol. May 15 2019;19(1):74. doi:10.1186/s12871-019-0735-5

33. Sadeghirad B, Dodsworth BT, Schmutz Gelsomino N, et al. Perioperative Factors Associated With Postoperative Delirium in Patients Undergoing Noncardiac Surgery: An Individual Patient Data Meta-Analysis. JAMA Netw Open. Oct 2 2023;6(10):e2337239. doi:10.1001/jamanetworkopen.2023.37239

34. Peden CJ, Miller TR, Deiner SG, Eckenhoff RG, Fleisher LA. Improving perioperative brain health: an expert consensus review of key actions for the perioperative care team. Br J Anaesth. Feb 2021;126(2):423–432. doi:10.1016/j.bja.2020.10.037

35. Jin Z, Hu J, Ma D. Postoperative delirium: perioperative assessment, risk reduction, and management. Br J Anaesth. Oct 2020;125(4):492–504. doi:10.1016/j.bja.2020.06.063

36. Menzenbach J, Kirfel A, Guttenthaler V, et al. PRe-Operative Prediction of postoperative DElirium by appropriate SCreening (PROPDESC) development and validation of a pragmatic POD risk screening score based on routine preoperative data. J Clin Anesth. Jun 2022;78:110684. doi:10.1016/j.jclinane.2022.110684

37. Bishara A, Chiu C, Whitlock EL, et al. Postoperative delirium prediction using machine learning models and preoperative electronic health record data. BMC Anesthesiol. Jan 3 2022;22(1):8. doi:10.1186/s12871-021-01543-y

38. Chen H, Mo L, Hu H, Ou Y, Luo J. Risk factors of postoperative delirium after cardiac surgery: a meta-analysis. J Cardiothorac Surg. Apr 26 2021;16(1):113. doi:10.1186/s13019-021-01496-w

39. Bramley P, McArthur K, Blayney A, McCullagh I. Risk factors for postoperative delirium: An umbrella review of systematic reviews. Int J Surg. Sep 2021;93:106063. doi:10.1016/j.ijsu.2021.106063

40. Sánchez A, Thomas C, Deeken F, et al. Patient safety, cost-effectiveness, and quality of life: reduction of delirium risk and postoperative cognitive dysfunction after elective procedures in older adults-study protocol for a stepped-wedge cluster randomized trial (PAWEL Study). Trials. Jan 21 2019;20(1):71. doi:10.1186/s13063-018-3148-8

41. Deeken F, Sánchez A, Rapp MA, et al. Outcomes of a Delirium Prevention Program in Older Persons After Elective Surgery: A Stepped-Wedge Cluster Randomized Clinical Trial. JAMA Surg. Feb 1 2022;157(2):e216370. doi:10.1001/jamasurg.2021.6370

42. Mathew G, Agha R, Albrecht J, et al. STROCSS 2021: Strengthening the reporting of cohort, cross-sectional and case-control studies in surgery. Int J Surg. Dec 2021;96:106165. doi:10.1016/j.ijsu.2021.106165

43. Jameson JL, Fauci AS, Kasper DL, Hauser SL, Longo DL, Loscalzo J. Editors. Harrison’s Principles of Internal Medicine, 20e. McGraw-Hill Education; 2018.

44. Chawla N, Bowyer K, Hall LO, Kegelmeyer WP. SMOTE: Synthetic Minority Over-sampling Technique. ArXiv. 2002;abs/1106.1813

45. Pedregosa F, Varoquaux G, Gramfort A, et al. Scikit-learn: Machine Learning in Python. Journal of Machine Learning Research. October 01, 2011 2011;12:2825–2830. doi:10.48550/arXiv.1201.0490

46. Chen T, Guestrin C. XGBoost: A Scalable Tree Boosting System. 2016:arXiv:1603.02754. doi:10.48550/arXiv.1603.02754 Accessed March 01, 2016. https://ui.adsabs.harvard.edu/abs/2016arXiv160302754C

47. Rutherford S, Barkema P, Tso IF, et al. Evidence for embracing normative modeling. eLife. 2023/03/13 2023;12:e85082. doi:10.7554/eLife.85082

48. Lundberg S, Lee S-I. A Unified Approach to Interpreting Model Predictions. 2017:arXiv:1705.07874. doi:10.48550/arXiv.1705.07874 Accessed May 01, 2017. https://ui.adsabs.harvard.edu/abs/2017arXiv170507874L

49. O’Neal JB, Billings FTt, Liu X, et al. Risk factors for delirium after cardiac surgery: a historical cohort study outlining the influence of cardiopulmonary bypass. Can J Anaesth. Nov 2017;64(11):1129–1137. Facteurs de risque de délirium après la circulation extracorporelle: une étude de cohorte historique décrivant l’influence de la circulation extracorporelle. doi:10.1007/s12630-017-0938-5

50. Levin P. Chapter 223 - Postoperative Delirium. In: Atlee JL, ed. Complications in Anesthesia (Second Edition). W.B. Saunders; 2007:888–889.

51. Kaźmierski J, Miler P, Pawlak A, et al. Lower Preoperative Verbal Memory Performance Is Associated with Delirium after Coronary Artery Bypass Graft Surgery: A Prospective Cohort Study. Arch Clin Neuropsychol. Jan 21 2023;38(1):49–56. doi:10.1093/arclin/acac064

52. Butz M, Meyer R, Gerriets T, et al. Increasing preoperative cognitive reserve to prevent postoperative delirium and postoperative cognitive decline in cardiac surgical patients (INCORE): Study protocol for a randomized clinical trial on cognitive training. Front Neurol. 2022;13:1040733. doi:10.3389/fneur.2022.1040733

53. Moller JT, Cluitmans P, Rasmussen LS, et al. Long-term postoperative cognitive dysfunction in the elderly ISPOCD1 study. ISPOCD investigators. International Study of Post-Operative Cognitive Dysfunction. Lancet. Mar 21 1998;351(9106):857–61. doi:10.1016/s0140-6736(97)07382-0

54. Weiss Y, Zac L, Refaeli E, et al. Preoperative Cognitive Impairment and Postoperative Delirium in Elderly Surgical Patients: A Retrospective Large Cohort Study (The CIPOD Study). Ann Surg. Jul 1 2023;278(1):59–64. doi:10.1097/sla.0000000000005657

55. Zhao J, Liang G, Hong K, et al. Risk factors for postoperative delirium following total hip or knee arthroplasty: A meta-analysis. Front Psychol. 2022;13:993136. doi:10.3389/fpsyg.2022.993136

56. Frei BW, Woodward KT, Zhang MY, et al. Considerations for Clock Drawing Scoring Systems in Perioperative Anesthesia Settings. Anesth Analg. May 2019;128(5):e61–e64. doi:10.1213/ane.0000000000004105

57. Fryer DV, Strümke I, Nguyen H. Shapley values for feature selection: The good, the bad, and the axioms. IEEE Access. 2021;PP:1–1.

58. Petch J, Di S, Nelson W. Opening the Black Box: The Promise and Limitations of Explainable Machine Learning in Cardiology. Can J Cardiol. Feb 2022;38(2):204–213. doi:10.1016/j.cjca.2021.09.004

59. Oh S, Park Y, Cho KJ, Kim SJ. Explainable Machine Learning Model for Glaucoma Diagnosis and Its Interpretation. Diagnostics (Basel). Mar 13 2021;11(3)doi:10.3390/diagnostics11030510

60. Amparore E, Perotti A, Bajardi P. To trust or not to trust an explanation: using LEAF to evaluate local linear XAI methods. PeerJ Comput Sci. 2021;7:e479. doi:10.7717/peerj-cs.479

61. Bordt S, von Luxburg U. From Shapley values to generalized additive models and back. PMLR; 2023:709–745.

62. Moon KJ, Son CS, Lee JH, Park M. The development of a web-based app employing machine learning for delirium prevention in long-term care facilities in South Korea. BMC Med Inform Decis Mak. Aug 17 2022;22(1):220. doi:10.1186/s12911-022-01966-8

