## Supplemental Tables and Figures for "Predicting Postoperative Delirium in Older Patients: a multicenter retrospective cohort study"

**Supplementary Digital Content**

**eFigure 1.** Model performances with four classifiers

**eFigure 2.** Receiver operating characteristic curves of models

**eFigure 3.** Pairwise comparisons of models with different features

**eFigure 4.** Pairwise comparisons of models with additional neuropsychological assessments

**eFigure 5.** Feature attribution for pre- and perioperative surgical features

**eFigure 6.** Feature attribution for preoperative neuropsychological assessments

**eFigure 7.** Sensitivity analysis of adding intervention allocation

**eFigure 8.** Sensitivity analysis of oversampling with the Synthetic Minority Oversampling Technique (SMOTE)

**eFigure 9.** Sensitivity analysis without outliers

**eTable 1.** Patient characteristics and information

**eTable 2.** Patient’s outcome after surgery

**eTable 3.** Information about missing values in data and outcome

**eTable 4.** Performance across all feature combinations, including intervention information, in various metrics.

**eTable 5.** Model performance in area under the receiver operating characteristic curve compared to random chance: 1000 permutation tests.

**eTable 6.** Model performance difference in area under the receiver operating characteristic curve: 1000 permutation tests.

**eTable 7.** Performance difference in the area under the receiver operating characteristic curve between models with additional intervention allocation information: 1000 permutation tests.

**eFigure 1.** Model performances with four classifiers

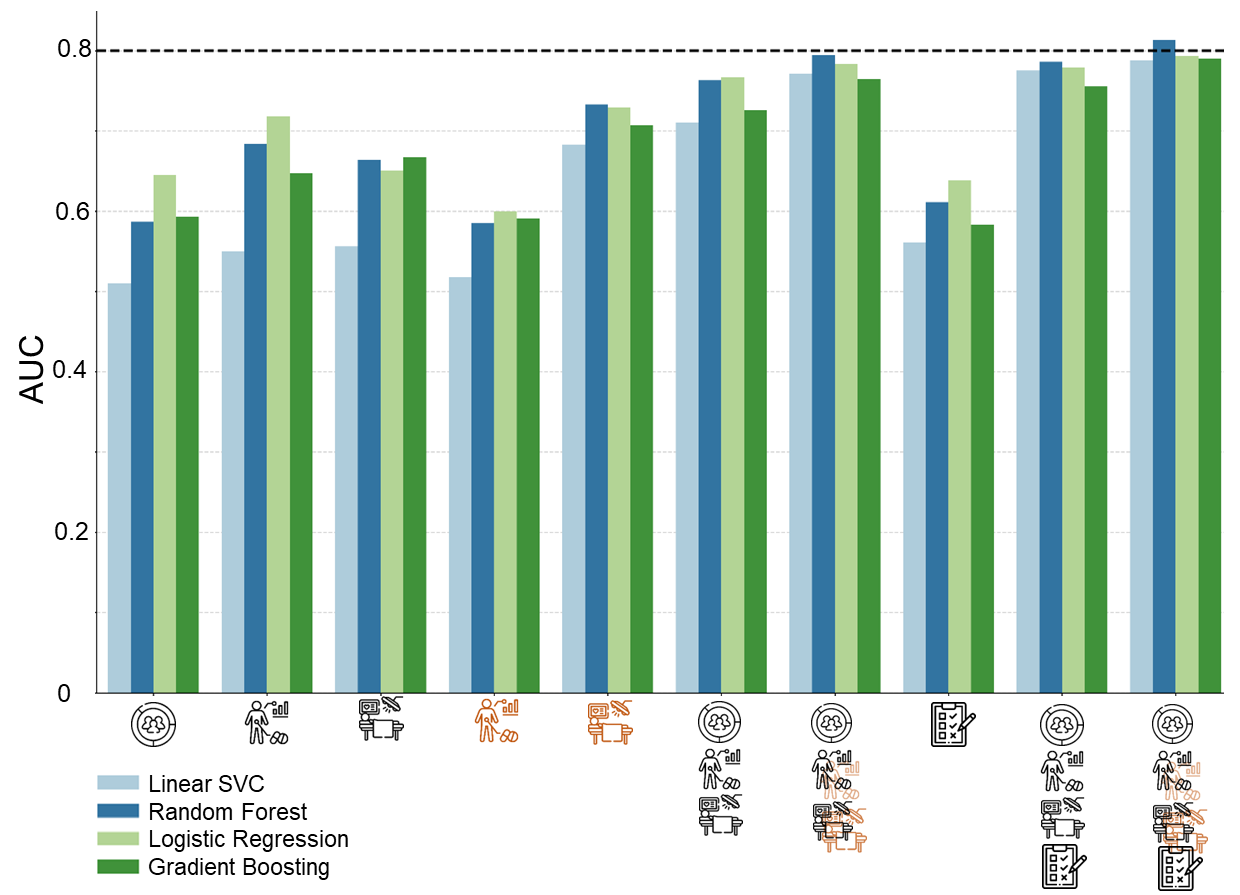

Comparable areas under the receiver operating characteristic curve (AUC) performance were observed across four classifiers. Notably, random forest algorithms exhibited slight superiority.

**eFigure 2.** Receiver operating characteristic curves

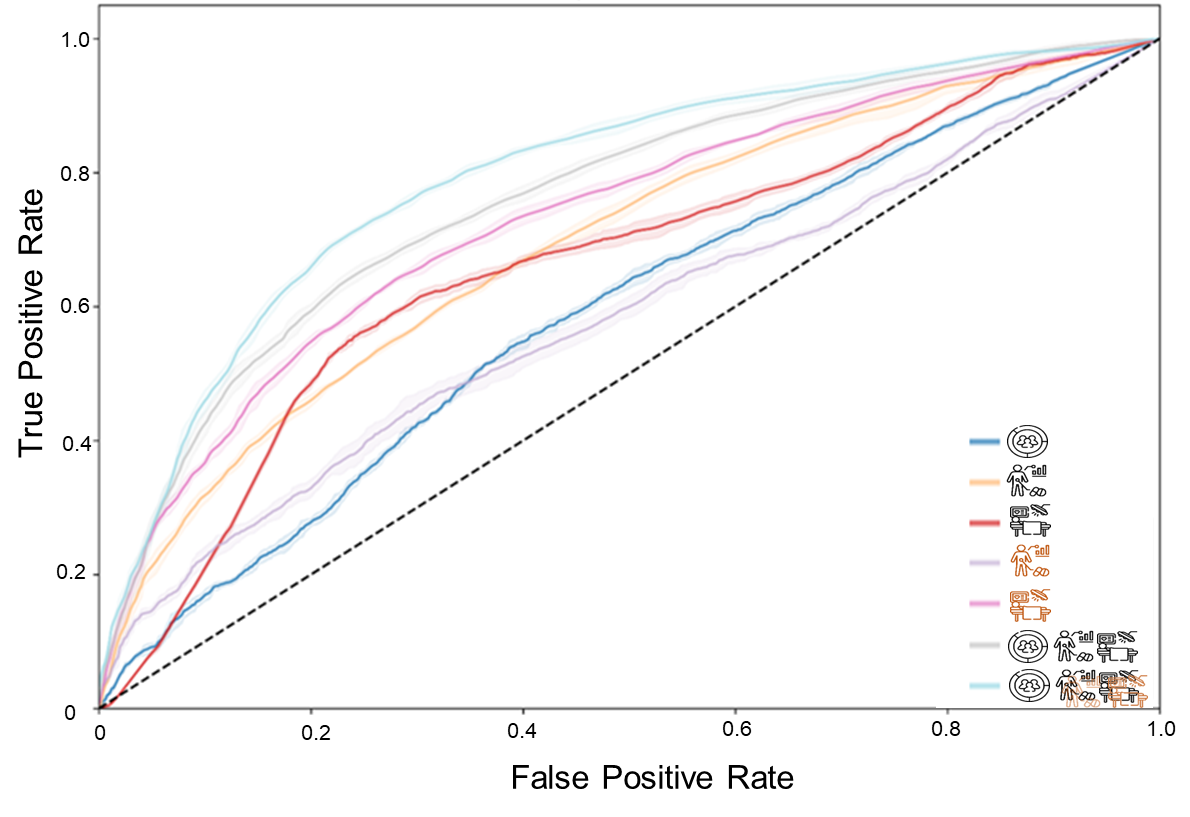

The receiver operating characteristic curve of models with combined and independent feature categories.

**eFigure 3.** Pairwise comparisons of models with different features

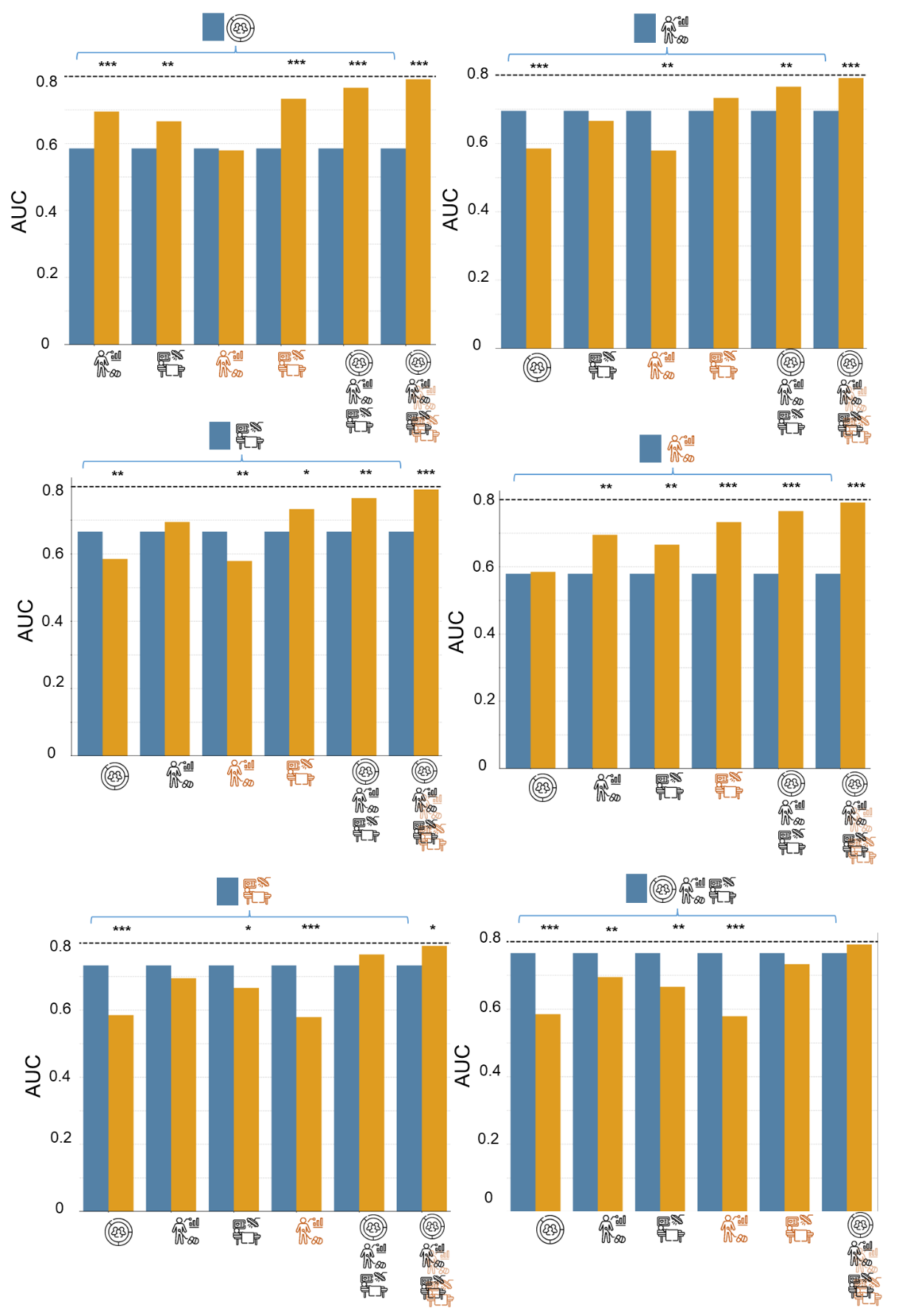

Comparisons of areas under the receiver operating characteristic curve (AUC) between models, with asterisks (*) indicating statistical significance levels for AUC differences based on 1000 permutations: **P* <.05, ***P* ≤.01, ****P* ≤.001. The blue bar represents the reference model, while the yellow bars represent the comparison models.

**eFigure 4.** Pairwise comparisons of models with additional neuropsychological assessments

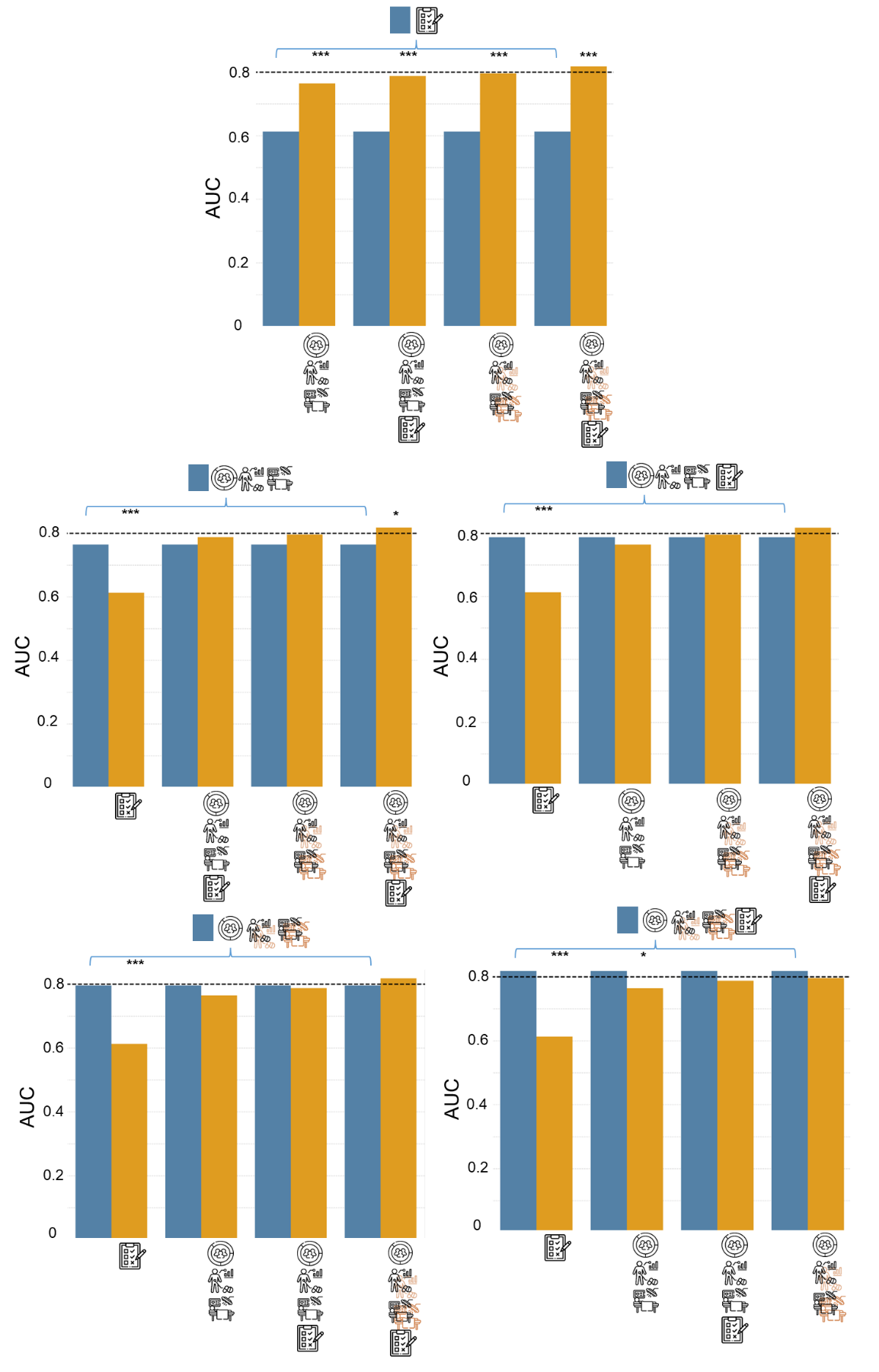

Comparisons of areas under the receiver operating characteristic curve (AUC) between models, with asterisks (*) indicating statistical significance levels for AUC differences based on 1000 permutations: **P* <.05, ***P* ≤.01, ****P* ≤.001. The blue bar represents the reference model, while the yellow bars represent the comparison models.

**eFigure 5.** Feature attribution for pre- and perioperative surgical features

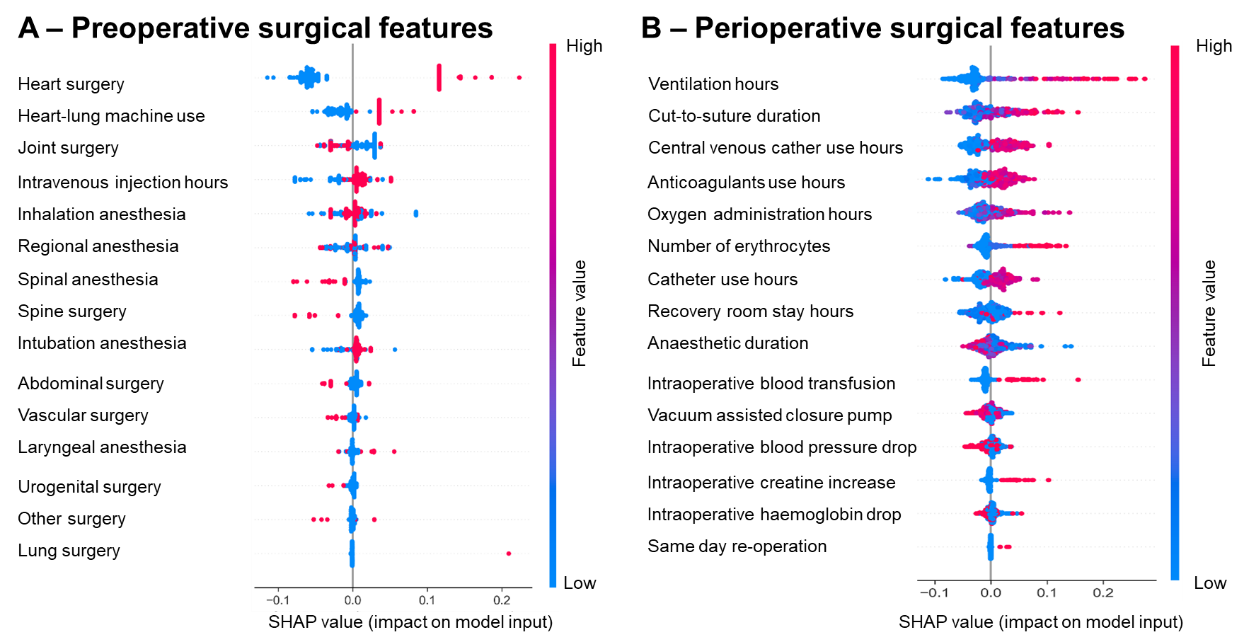

(A) SHapley Additive exPlanations (SHAP)-based preoperative surgical feature attribution reveals higher postoperative delirium (POD) risk in patients undergoing heart surgeries with heart-lung machines. (B) SHAP analysis assigns increased POD risk to patients with longer operation/anesthesia durations and prolonged durations of surgical equipment usage. The dot color indicates feature values (blue for low, red for high), while the horizontal position reflects feature impact on model output (left for negative, right for positive).

**eFigure 6.** Feature attribution for preoperative neuropsychological assessments

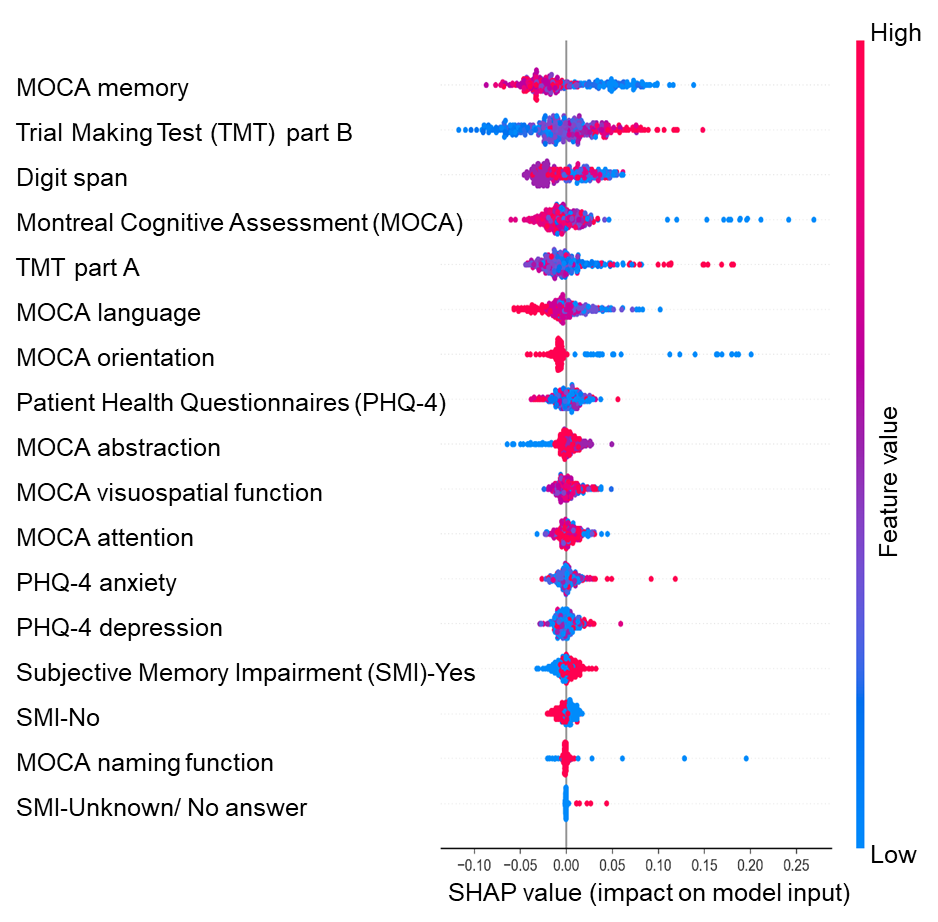

SHapley Additive Explanations (SHAP) analysis reveals that lower scores in the Montreal Cognitive Assessment (MOCA) memory subdomain and longer Trail Making Test (TMT) part B completion times are associated with an increased risk of postoperative delirium (POD). Feature values are represented by dot color (blue for low, red for high), and feature impact on model output is shown through horizontal positioning (left for negative, right for positive).

**eFigure 7.** Sensitivity analysis of adding intervention allocation

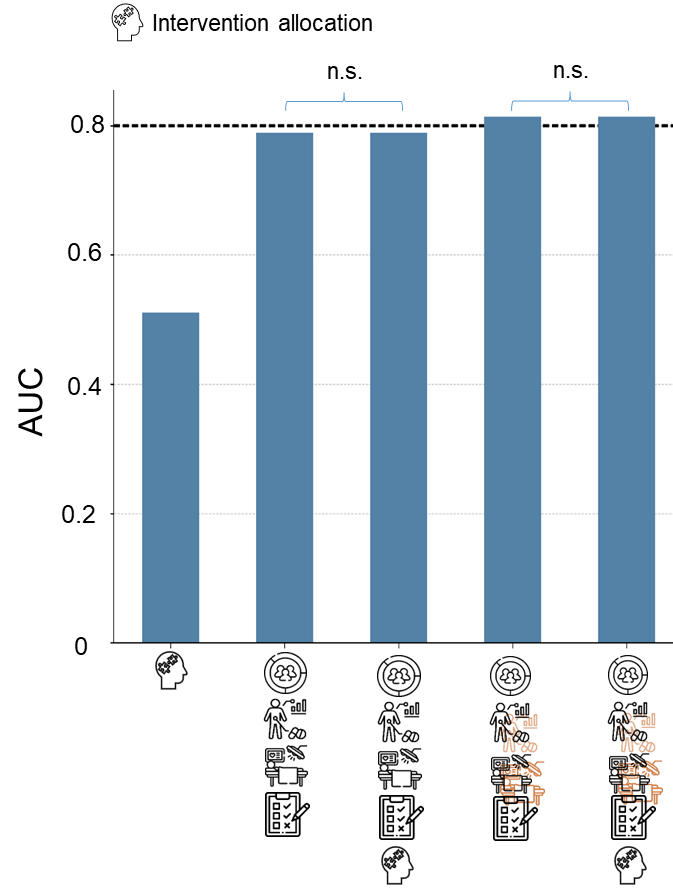

This sensitivity analysis demonstrates that intervention allocation does not impact model performance. n.s. indicates no statistically significant difference between models. AUC shows areas under the receiver operating characteristic curve.

**eFigure 8.** Sensitivity analysis of oversampling with the Synthetic Minority Oversampling Technique (SMOTE)

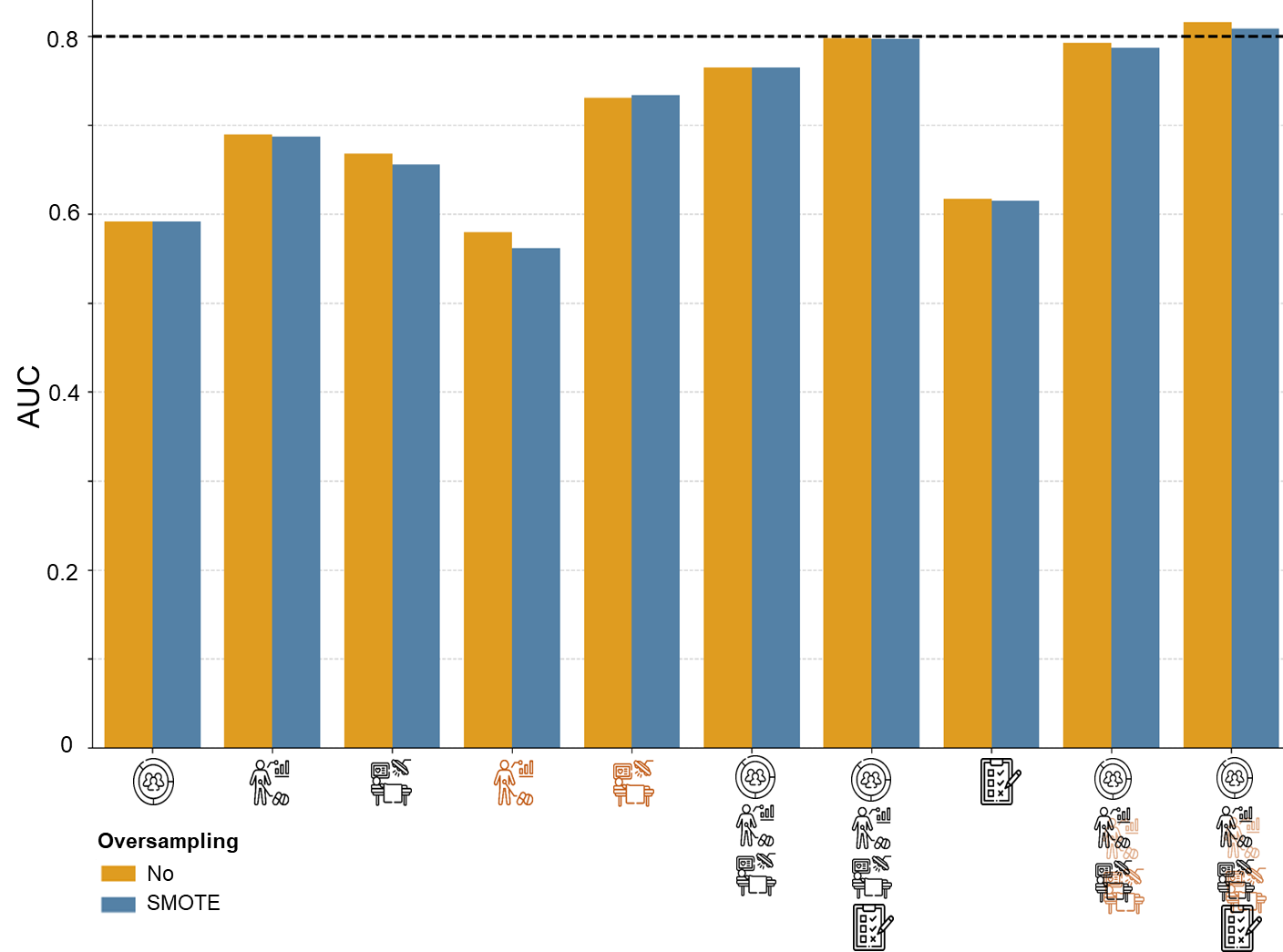

The sensitivity analysis for potential imbalance by the SMOTE highlights that there is no discernible distinction in the areas under the receiver operating characteristic curve (AUC) between models with and without oversampling.

**eFigure 9.** Sensitivity analysis without outliers

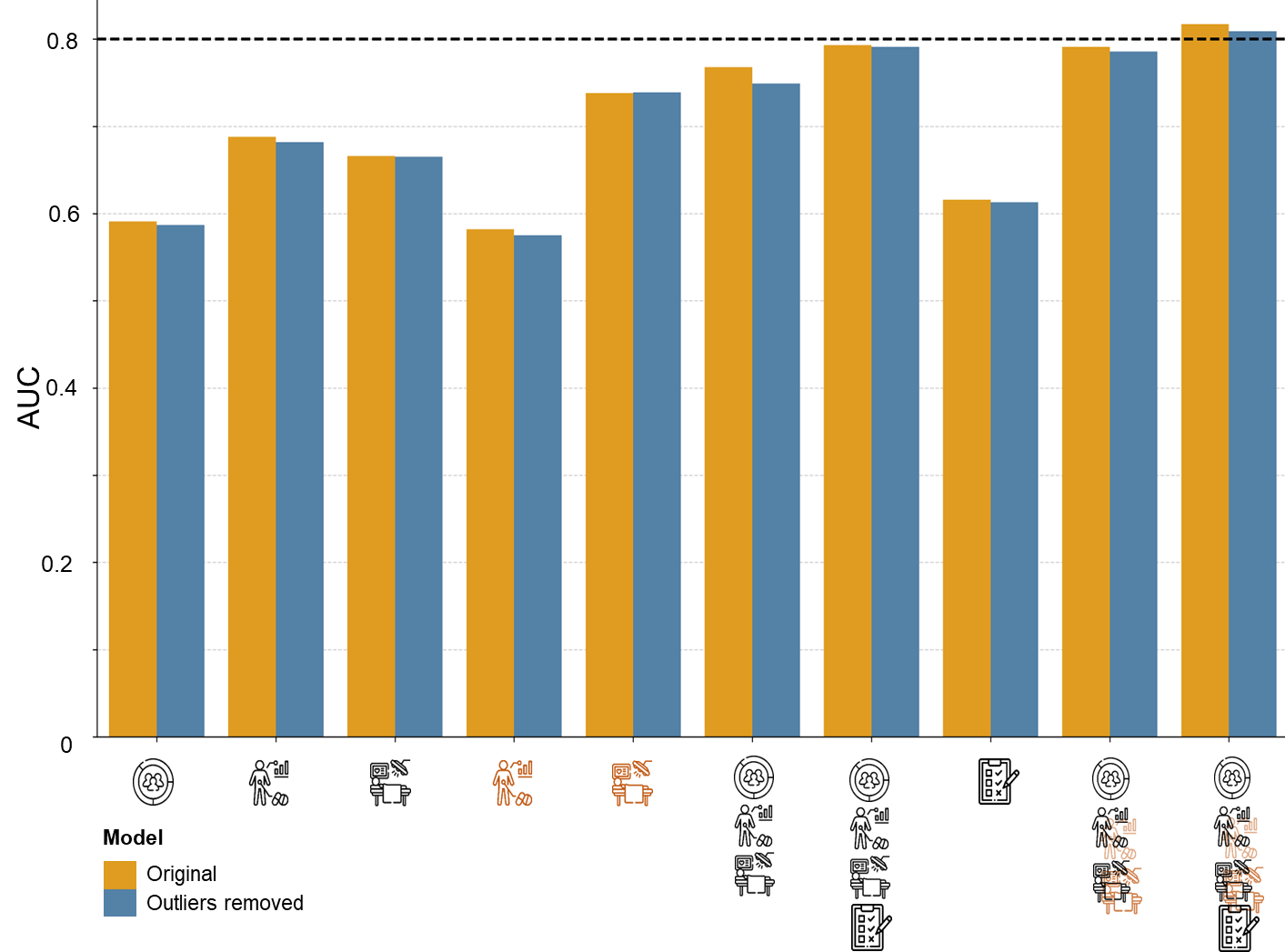

The sensitivity analysis for potential outliers reveals that there is no discernible distinction in the areas under the receiver operating characteristic curve (AUC) between models with and without outliers.
**eTable 1.** Patient characteristics and information

| Demographics | Delirium (N=375) | Non-Delirium (N=1249) | *P* |
| --- | --- | --- | --- |
| Male (%) | 237 (63.2%) | 613 (49.1%) | <.001** |
| Mean ages (SD)^+^ | 78.34 (4.97) | 77.74 (4.86) | .195 |
| Number of participants in each location | 38:47:74:  102:114 | 235:286:256:  238:234 | <.001** |
| Years of education in total (SD) | 12.6 (2.87) | 12.84 (2.9) | .134 |
| Alcohol consumption per month (SD) | .03 (.18) | .02 (.15) | .430 |
| Cigarette consumption per day (SD) ^+^ | .59 (2.67) | .58 (2.89) | .874 |
| Alone living (%) | 113 (30.1%) | 362 (29.0%) | .715 |
| Received non-pharmacological interventions for delirium (%) | 163 (43.5%) | 606 (48.5%) | .097 |
| Cognitive information and psychological screens | **Delirium (N=375)** | **Non-Delirium (N=1249)** | ***P*** |
| Montreal Cognitive Assessment (MoCA) sum score at admission (SD) | 20.97 (4.63) | 22.99 (3.41) | <.001** |
| MoCA Attention at admission (SD) | 4.94 (1.31) | 5.25 (.99) | <.001** |
| MoCA Memory at admission (SD) | 1.74 (1.66) | 2.55 (1.65) | <.001** |
| MoCA Orientation at admission (SD) | 5.60 (0.97) | 5.89 (0.41) | <.001** |
| MoCA Visuospatial/Executive at admission (SD) | 2.91 (1.44) | 3.21 (1.31) | <.001** |
| MoCA Naming at admission (SD) | 2.86 (0.48) | 2.92 (0.32) | .025* |
| MoCA Language at admission (SD) | 1.57 (0.94) | 1.84 (0.91) | <.001** |
| MoCA Abstraction at admission (SD) | 1.44 (0.69) | 1.53 (0.67) | .034* |
| Trial Making Test part A (TMT-A) at admission (SD) | 74.01 (39.5) | 62.33 (28.2) | <.001** |
| Trial Making Test part B (TMT-B) at admission (SD) | 183.6 (75.2) | 154.3 (68.2) | <.001** |
| Digit span (length) at admission (SD) | 3.71 (1.29) | 4.0 (1.19) | <.001** |
| Patient Health Questionnaire-4 (PHQ-4) at admission (median, SD)^+^ | 1 (2.78) | 1 (2.61) | .718 |
| PHQ-4 anxiety rating at admission (median, SD)^+^ | 1 (1.49) | 1 (1.39) | .417 |
| PHQ-4 depression rating at admission (median, SD)^+^ | 1 (1.66) | 1 (1.58) | .289 |
| Subjective memory impairment (SMI) at admission (median, SD)^+^ | 1 (.55) | 1 (.53) | .121 |
| Clinical information | **Delirium (N=375)** | **Non-Delirium (N=1249)** | ***P*** |
| Hemoglobin level (g/dl) at admission (SD) | 13.42 (6.53) | 13.65 (7.33) | .572 |
| Sodium level (mmol/l) at admission (SD) | 140.1 (2.86) | 139.9 (3.00) | .354 |
| C-reactive protein (mg/l) at admission (SD) | 9.21 (18.98) | 8.44 (19.97) | .530 |
| Renal failure (creatinine clearance in mg/dl) at admission (SD) | 64.99 (24.4) | 69.00 (30.7) | .010* |
| Polypharmacy (number of medicines taken) at admission (SD)^+^ | 7.28 (3.71) | 6.03 (3.65) | .012* |
| Dose (mg) of preoperative Benzodiazepines use (SD)^+^ | 7.90 (14.42) | 3.55 (13.24) | <.001** |
| Dose (mg) of preoperative Neuroleptics use (SD)^+^ | 1.05 (10.3) | .14 (2.47) | 0.617 |
| Dose (mg) of preoperative Opiates use (SD) | 0.83 (11.57) | 2.33 (21.3) | .081 |
| Dose (mg) of preoperative Propofol use (SD) | 2.57 (19.3) | 9.78 (105.5) | .023* |
| Mini Nutritional Assessment (MNA) sum score at admission (SD) | 11.08 (2.71) | 11.43 (2.58) | .026* |
| Multimorbidity (number of diseases reported) at admission (SD)^+^ | .96 (.66) | .78 (.67) | .113 |
| ASA Physical Status Classification System (median) at admission (SD) | 3 (.60) | 3 (.63) | <.001** |
| Charlson Comorbidity Index at admission (SD)^+^ | 2.53 (2.23) | 2.21 (2.25) | .255 |
| Barthel Index sum score at admission (SD) | 91.08 (19.4) | 93.66 (14.6) | .02* |
| Clinical Frailty Scale (median) at admission (SD) | 3 (1.41) | 3 (1.33) | .005* |
| Auditory impairment at admission (%) | 251 (68.6%) | 674 (56.5%) | .027* |
| Visual impairment at admission (%) | 74 (20.6%) | 229 (19.7%) | .764 |
| Any sensory impairment at admission (%) | 233 (65.3%) | 650 (56.3%) | .025* |
| Preexisting dementia (%) | 22 (5.87%) | 7 (.56%) | <.001** |
| Previous delirium history (%) | 38 (10.38%) | 100 (8.17%) | .225 |
| Surgery | **Delirium (N=375)** | **Non-Delirium (N=1249)** | ***P*** |
| Minutes of cut-to-suture time (SD)^+^ | 191.3 (95.5) | 131.5 (75.4) | <.001** |
| Minutes of anesthetic duration (SD) | 252.9 (104) | 201.0 (84.5) | <.001** |
| Cardio-pulmonary bypass (%) | 176 (47.1%) | 220 (17.8%) | <.001** |
| Types of anesthesia (Intubation: Laryngeal: Spinal) | 318:14:31 | 907:78:207 | <.001** |
| Inhalation anesthesia (%) | 284 (75.7%) | 751 (60.1%) | <.001** |
| Regional anesthesia (%) | 87 (23.2%) | 469 (37.6%) | <.001** |
| Types of surgery  (Joint: Spine: Vessels: Heart: Lung: Abdominal: Urogenital: Others) | 100:11:22:  206:1:21:7:7 | 574:100:126:  264:1:108:49:27 | <.001** |
| Dose (mg) of intraoperative Benzodiazepines use (SD)^+^ | 1.75 (4.85) | 1.41 (14.21) | .952 |
| Dose (mg) of intraoperative Neuroleptics use (SD)^+^ | .16 (2.02) | .08 (2.14) | .617 |
| Dose (mg) of intraoperative Opiates use (SD) | 9.65 (81.3) | 10.82 (208) | .872 |
| Dose (mg) of intraoperative Propofol use (SD) | 522 (675.5) | 297 (471.8) | <.001** |
| Number of erythrocytes during surgery (SD) | .91 (1.68) | .28 (.90) | <.001** |
| Blood pressure drop during surgery (%) | 125 (33.6%) | 488 (39.5%) | .046* |
| Blood transfusion during surgery (%) | 122 (32.8%) | 133 (10.7%) | <.001** |
| Drop in hemoglobin value >3g/dl during surgery (%) | 196 (53.3%) | 449 (37.5%) | <.001** |
| Use of a heart-lung machine during surgery (%) | 176 (47.1%) | 220 (17.8%) | <.001** |
| Increase in creatine >0.3mg/dl during surgery (%) | 49 (13.5%) | 59 (4.8%) | <.001** |
| Another operation (re-operation) on the same day (%) | 11 (2.93%) | 11 (.88%) | .006* |
| Hours of anticoagulants (AVK) use after surgery (median, SD) | 33 (39.54) | 24 (23.24) | <.001** |
| Hours of catheter use after surgery (median, SD) | 34 (34.99) | 29 (27.63) | <.001** |
| Hours of O2 administration after surgery (median, SD) | 20 (37.0) | 15 (23.46) | .001** |
| Hours in the recovery room after surgery (median, SD) | 0 (14.71) | 1 (6.05) | .787 |
| Hours of ventilation after surgery (median, SD) | 3 (12.51) | 0 (9.01) | <.001** |
| Hours of Vacuum Assisted Closure pump after surgery (median, SD) | 34.0 (18.21) | 33.0 (21.45) | <.001** |
| Hours of Central Venous Catheter use after surgery (median, SD) | 33.0 (24.69) | 6.0 (28.43) | <.001** |

^+^: nonparametric Mann-Whitney U tests for continuous/discrete variables without normality or ordinal variables; **P* <.05, ***P* ≤.001
**eTable 2.** Patient’s outcome after surgery

| Outcome | Delirium  (N=375) | Non-Delirium  (N=1249) | *P* |
| --- | --- | --- | --- |
| Attention deficit in CAM at discharge (%) | 42 (11.38%) | 52 (4.52%) | <.001** |
| Disorganized thinking rating in CAM at discharge (SD) | 0 (.49) | 0 (.38) | .005* |
| Hyperactivity in CAM at discharge (%) | 4 (1.08%) | 2 (0.16%) | .040* |
| Motor problems in CAM at discharge (%) | 7 (1.9%) | 11 (.89%) | .120 |
| Drowsiness in CAM at discharge (%) | 5 (1.35%) | 3 (0.24%) | .025* |
| Barthel Index sum score at discharge (SD) | 74.82 (26.3) | 88.4 (15.6) | <.001** |
| MoCA sum score at discharge (SD) | 20.01 (4.69) | 22.65 (3.57) | <.001** |
| MoCA Attention at discharge (SD) | 4.67 (1.45) | 5.17 (1.05) | <.001** |
| MoCA Memory at discharge (SD) | 1.62 (1.58) | 2.35 (1.63) | <.001** |
| MoCA Orientation at discharge (SD) | 5.42 (1.06) | 5.8 (0.54) | <.001** |
| MoCA Visuospatial/Executive at discharge (SD) | 2.89 (1.45) | 3.37 (1.30) | <.001** |
| MoCA Naming at discharge (SD) | 2.58 (0.64) | 2.72 (0.55) | <.001** |
| MoCA Language at discharge (SD) | 1.14 (0.98) | 1.62 (1.04) | <.001** |
| MoCA Abstraction at discharge (SD) | 1.72 (0.57) | 1.86 (0.42) | <.001** |
| TMT-A at discharge (SD) | 81.39 (43.0) | 64.27 (31.7) | <.001** |
| TMT-B at discharge (SD) | 198.5 (77.2) | 158.0 (71.4) | <.001** |
| Digit span (length) at discharge (SD) | 3.49 (1.33) | 3.96 (1.20) | <.001** |
| Days in Intermediate Care Unit stay after surgery (SD)^+^ | .77 (1.52) | .39 (.91) | .637 |
| Days of Intensive Care Unit stay after surgery (SD)^+^ | 2.60 (2.70) | .68 (1.19) | <.001** |
| Days of hospitalization (SD)^+^ | 8.66 (2.06) | 8.02 (2.31) | <.001** |

^+^: nonparametric Mann-Whitney U tests for continuous/discrete variables without normality or ordinal variables; *: P<.05; **: P<.001

**eTable 3**. Information about missing values in data and outcome

| Demographics | Delirium (N=375) | Non-Delirium (N=1249) | All subjects  (N=1624) |
| --- | --- | --- | --- |
| Sex (%) | 0 (0%) | 0 (0%) | 0 (0%) |
| Mean ages (%) | 0 (0%) | 0 (0%) | 0 (0%) |
| Location (%) | 0 (0%) | 0 (0%) | 0 (0%) |
| Years of education in total (%) | 2 (.53%) | 3 (.24%) | 5 (.31%) |
| Alcohol consumption per month (%) | 2 (.53%) | 8 (.64%) | 10 (.62%) |
| Cigarette consumption per day (SD) | 0 (0%) | 0 (0%) | 0 (0%) |
| Alone living (%) | 0 (0%) | 0 (0%) | 0 (0%) |
| Received non-pharmacological interventions for delirium (%) | 0 (0%) | 0 (0%) | 0 (0%) |
| Cognitive information and psychological screens | **Delirium (N=375)** | **Non-Delirium (N=1249)** | **All subjects**  **(N=1624)** |
| Montreal Cognitive Assessment (MoCA) sum score at admission (%) | 14 (3.73%) | 34 (2.72%) | 48 (2.96%) |
| MoCA Attention at admission (%) | 5 (1.33%) | 12 (.96%) | 17 (1.05%) |
| MoCA Memory at admission (%) | 5 (1.33%) | 14 (1.12%) | 19 (1.17%) |
| MoCA Orientation at admission (%) | 5 (1.33%) | 11 (.88%) | 16 (.99%) |
| MoCA Visuospatial/Executive at admission (%) | 12 (3.2%) | 29 (2.32%) | 41 (2.52%) |
| MoCA Naming at admission (%) | 4 (1.07%) | 9 (.72%) | 13 (.8%) |
| MoCA Language at admission (%) | 5 (1.33%) | 13 (1.04%) | 18 (1.11%) |
| MoCA Abstraction at admission (%) | 5 (1.33%) | 14 (1.12%) | 19 (1.17%) |
| Trial Making Test part A (TMT-A) at admission (%) | 53 (14.13%) | 158 (12.65%) | 211 (12.99%) |
| Trial Making Test part B (TMT-B) at admission (%) | 81 (21.6%) | 223 (17.85%) | 304 (18.72%) |
| Digit span (length) at admission (%) | 56 (14.93%) | 152 (12.17%) | 208 (12.81%) |
| Patient Health Questionnaire-4 (PHQ-4) at admission (%) | 17 (4.53%) | 57 (4.56%) | 74 (4.56%) |
| PHQ-4 anxiety rating at admission (%) | 17 (4.53%) | 53 (4.24%) | 70 (4.31%) |
| PHQ-4 depression rating at admission (%) | 17 (4.53%) | 56 (4.48%) | 73 (4.5%) |
| Subjective memory impairment (SMI) at admission (%) | 1 (.27%) | 4 (0.32%) | 5 (.31%) |
| Clinical information | **Delirium (N=375)** | **Non-Delirium (N=1249)** | **All subjects**  **(N=1624)** |
| Hemoglobin level (g/dl) at admission (%) | 6 (1.6%) | 30 (2.4%) | 36 (2.22%) |
| Sodium level (mmol/l) at admission (%) | 8 (2.13%) | 36 (2.88%) | 44 (2.71%) |
| C-reactive protein (mg/l) at admission (%) | 53 (14.13%) | 246 (19.7%) | 299 (18.41%) |
| Renal failure (creatinine clearance in mg/dl) at admission (%) | 11 (2.93%) | 43 (3.44%) | 54 (3.33%) |
| Polypharmacy (number of medicines taken) at admission (%) | 0 (0%) | 0 (0%) | 0 (0%) |
| Dose (mg) of preoperative Benzodiazepines use (%) | 0 (0%) | 3 (0.24%) | 3 (.18%) |
| Dose (mg) of preoperative Neuroleptics use (%) | 0 (0%) | 4 (0.32%) | 4 (.25%) |
| Dose (mg) of preoperative Opiates use (%) | 2 (0.53%) | 24 (1.92%) | 26 (1.6%) |
| Dose (mg) of preoperative Propofol use (%) | 5 (1.33%) | 18 (1.44%) | 23 (1.42%) |
| Mini Nutritional Assessment (MNA) sum score at admission (%) | 9 (2.4%) | 31 (2.48%) | 40 (2.46%) |
| Multimorbidity ( number of diseases reported) at admission (%) | 0 (0%) | 0 (0%) | 0 (0%) |
| ASA Physical Status Classification System (median) at admission (%) | 7 (1.87%) | 33 (2.64%) | 40 (2.46%) |
| Charlson Comorbidity Index at admission (%) | 0 (0%) | 0 (0%) | 0 (0%) |
| Barthel Index sum score at admission (%) | 10 (2.67%) | 37 (2.96%) | 47 (2.89%) |
| Clinical Fragility Scale (median) at admission (%) | 2 (.53%) | 19 (1.52%) | 21 (1.29%) |
| Auditory impairment at admission (%) | 9 (2.4%) | 57 (4.56%) | 66 (4.06%) |
| Visual impairment at admission (%) | 16 (4.27%) | 87 (6.97%) | 103 (6.34%) |
| Sensory impairment at admission (%) | 18 (4.8%) | 95 (7.61%) | 113 (6.96%) |
| Preexisting dementia (%) | 0 (0%) | 0 (0%) | 0 (0%) |
| Previous delirium history (%) | 9 (2.4%) | 25 (2%) | 34 (2.09%) |
| Surgery | **Delirium (N=375)** | **Non-Delirium (N=1249)** | **All subjects**  **(N=1624)** |
| Minutes of cut-to-suture time (%) | 1 (.27%) | 0 (0%) | 1 (.06%) |
| Minutes of anesthetic duration (%) | 79 (21.01%) | 126 (10.09%) | 205 (12.62%) |
| Cardio-pulmonary bypass (%) | 1 (.27%) | 11 (.88%) | 12 (.74%) |
| Types of anesthesia (%) | 12 (3.2%) | 57 (4.56%) | 69 (4.25%) |
| Inhalation anesthesia (%) | 0 (0%) | 0 (0%) | 0 (0%) |
| Regional anesthesia (%) | 0 (0%) | 0 (0%) | 0 (0%) |
| Types of surgery (%) | 0 (0%) | 0 (0%) | 0 (0%) |
| Dose (mg) of intraoperative Benzodiazepines use (%) | 0 (0%) | 3 (.24%) | 3 (.18%) |
| Dose (mg) of intraoperative Neuroleptics use (%) | 0 (0%) | 4 (.32%) | 4 (.25%) |
| Dose (mg) of intraoperative Opiates use (%) | 2 (.53%) | 19 (1.52%) | 21 (1.29%) |
| Dose (mg) of intraoperative Propofol use (%) | 2 (.53%) | 5 (.4%) | 7 (.43%) |
| Number of erythrocytes during surgery (%) | 26 (6.93%) | 127 (10.17%) | 153 (9.42%) |
| Blood pressure drop during surgery (%) | 3 (.8%) | 14 (1.12%) | 17 (1.05%) |
| Blood transfusion during surgery (%) | 3 (.8%) | 10 (.8%) | 13 (.8%) |
| Drop in hemoglobin value >3g/dl during surgery (%) | 7 (1.87%) | 50 (4%) | 57 (3.51%) |
| Use of a heart-lung machine during surgery (%) | 1 (.27%) | 11 (.88%) | 12 (.74%) |
| Increase in creatine >0.3mg/dl during surgery (%) | 12 (3.2%) | 77 (6.16%) | 89 (5.48%) |
| Another operation (re-operation) on the same day (%) | 0 (0%) | 0 (0%) | 0 (0%) |
| Hours of anticoagulants (AVK) use after surgery (%) | 1 (.27%) | 17 (1.36%) | 18 (1.11%) |
| Hours of catheter use after surgery (%) | 1 (.27%) | 13 (1.04%) | 14 (.86%) |
| Hours of O2 administration after surgery (%) | 4 (1.07%) | 17 (1.36%) | 21 (1.29%) |
| Hours in the recovery room after surgery (%) | 2 (.53%) | 20 (1.6%) | 22 (1.35%) |
| Hours of ventilation after surgery (%) | 2 (.53%) | 13 (1.04%) | 15 (.92%) |
| Hours of Vacuum Assisted Closure pump after surgery (%) | 13 (1.04%) | 14 (1.12%) | 27 (1.66%) |
| Hours of Central Venous Catheter use after surgery (%) | 4 (1.12%) | 20 (1.6%) | 24 (1.48%) |
| Outcome | **Delirium (N=375)** | **Non-Delirium (N=1249)** | **All subjects**  **(N=1624)** |
| Post-operative delirium (%) | 0 (0%) | 0 (0%) | 0 (0%) |

**eTable 4.** Performance across all feature combinations, including intervention information, in various metrics.

| Model | ROC | PRC | Sen | Spe | Prec | Rec | BA |
| --- | --- | --- | --- | --- | --- | --- | --- |
| Preoperative: Demographic | 0.59 | 0.30 | 21.9 | 86.2 | 32.2 | 21.9 | 54.0 |
| Preoperative: Clinical | 0.70 | 0.42 | 16.3 | 95.9 | 54.5 | 16.3 | 56.1 |
| Preoperative: Surgical | 0.67 | 0.36 | 0.5 | 99.2 | 16.7 | 0.5 | 49.9 |
| Perioperative: Clinical | 0.58 | 0.35 | 21.9 | 89.0 | 37.4 | 21.9 | 55.5 |
| Perioperative: Surgical | 0.74 | 0.49 | 30.7 | 92.4 | 54.8 | 30.7 | 61.5 |
| Preoperative: Demographic, Clinical, Surgical | 0.76 | 0.52 | 30.7 | 94.3 | 61.8 | 30.7 | 62.5 |
| Preoperative: Demographic, Clinical, Surgical and  Perioperative: Clinical, Surgical | 0.80 | 0.55 | 32.8 | 94.1 | 62.4 | 32.8 | 63.4 |
| Preoperative: Neuropsychological | 0.61 | 0.35 | 10.9 | 96.5 | 48.2 | 10.9 | 53.7 |
| Preoperative: Demographic, Clinical, Surgical, Neuropsychological | 0.79 | 0.54 | 31.2 | 94.1 | 61.3 | 31.2 | 62.6 |
| Preoperative: Demographic, Clinical, Surgical, Neuropsychological, Intervention group | 0.79 | 0.54 | 28.8 | 95.6 | 66.3 | 28.8 | 62.2 |
| Preoperative: Demographic, Clinical, Surgical, Neuropsychological and Perioperative: Clinical, Surgical | 0.82 | 0.57 | 32.8 | 95.0 | 66.1 | 32.8 | 63.9 |
| Preoperative: Demographic, Clinical, Surgical, Neuropsychological, Intervention allocation | 0.79 | 0.56 | 27.2 | 94.6 | 60.4 | 27.2 | 60.9 |
| Preoperative: Demographic, Clinical, Surgical, Neuropsychological, Intervention allocation and Perioperative: Clinical, Surgical | 0.82 | 0.58 | 34.9 | 95.2 | 68.6 | 34.9 | 65.1 |

ROC = area under the receiver operating characteristic curve; PRC= area under the precision-recall curve; Sen= sensitivity; Spe=specificity; Prec= precision; Rec= recall; BA= balanced accuracy.

**eTable 5.** Model performance in area under the receiver operating characteristic curve compared to random chance: 1000 permutation tests.

| Model | *P* |
| --- | --- |
| Preoperative: Demographic | 0.001998 |
| Preoperative: Clinical | 0.000999 |
| Preoperative: Surgical | 0.000999 |
| Perioperative: Clinical | 0.001998 |
| Perioperative: Surgical | 0.000999 |
| Preoperative: Demographic, Clinical, Surgical | 0.000999 |
| Preoperative: Demographic, Clinical, Surgical and  Perioperative: Clinical, Surgical | 0.000999 |
| Preoperative: Neuropsychological | 0.000999 |
| Preoperative: Demographic, Clinical, Surgical, Neuropsychological | 0.000999 |
| Preoperative: Demographic, Clinical, Surgical, Neuropsychological and Perioperative: Clinical, Surgical | 0.000999 |
| Preoperative: Intervention allocation | 0.135864 |
| Preoperative: Demographic, Clinical, Surgical, Neuropsychological, Intervention allocation | 0.000999 |
| Preoperative: Demographic, Clinical, Surgical, Neuropsychological, Intervention allocation and  Perioperative: Clinical, Surgical | 0.000999 |

**eTable 6.** Model performance difference in area under the receiver operating characteristic curve: 1000 permutation tests.

| *P* | 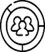 | 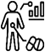 | 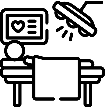 | 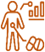 | 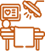 | 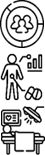 | 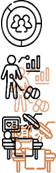 | 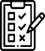 | 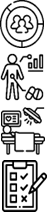 | 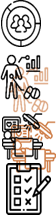 |
| --- | --- | --- | --- | --- | --- | --- | --- | --- | --- | --- |
| 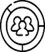 | 1.0000 | 0.0020 | 0.0210 | 0.7433 | 0.0010 | 0.0010 | 0.0010 | 0.5724 | 0.0010 | 0.0010 |
| 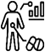 | 0.0020 | 1.0000 | 0.4046 | 0.0020 | 0.0939 | 0.0010 | 0.0010 | 0.0100 | 0.0010 | 0.0010 |
| 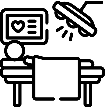 | 0.0210 | 0.4046 | 1.0000 | 0.0050 | 0.0180 | 0.0010 | 0.0010 | 0.0799 | 0.0010 | 0.0010 |
| 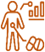 | 0.7433 | 0.0020 | 0.0050 | 1.0000 | 0.0010 | 0.0010 | 0.0010 | 0.3437 | 0.0010 | 0.0010 |
| 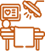 | 0.0010 | 0.0939 | 0.0180 | 0.0010 | 1.0000 | 0.4196 | 0.0180 | 0.0010 | 0.1319 | 0.0100 |
| 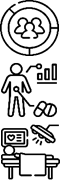 | 0.0010 | 0.0010 | 0.0010 | 0.0010 | 0.4196 | 1.0000 | 0.1209 | 0.0010 | 0.3237 | 0.0440 |
| 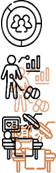 | 0.0010 | 0.0010 | 0.0010 | 0.0010 | 0.0180 | 0.1209 | 1.0000 | 0.0010 | 0.6384 | 0.4446 |
| 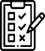 | 0.5724 | 0.0100 | 0.0799 | 0.3437 | 0.0010 | 0.0010 | 0.0010 | 1.0000 | 0.0010 | 0.0010 |
| 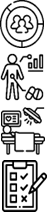 | 0.0010 | 0.0010 | 0.0010 | 0.0010 | 0.1319 | 0.3237 | 0.6384 | 0.0010 | 1.0000 | 0.1928 |
| 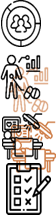 | 0.0010 | 0.0010 | 0.0010 | 0.0010 | 0.0100 | 0.0440 | 0.4446 | 0.0010 | 0.1928 | 1.0000 |

List of icons:

| Preoperative: Demographic | 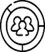 |
| --- | --- |
| Preoperative: Clinical | 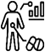 |
| Preoperative: Surgical | 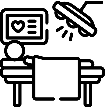 |
| Perioperative: Clinical | 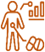 |
| Perioperative: Surgical | 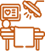 |
| Preoperative: Demographic, Clinical, Surgical | 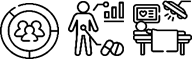 |
| Preoperative: Demographic, Clinical, Surgical and Perioperative: Clinical, Surgical | 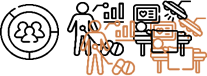 |
| Preoperative: Neuropsychological | 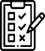 |
| Preoperative: Demographic, Clinical, Surgical, Neuropsychological | 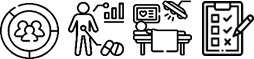 |
| Preoperative: Demographic, Clinical, Surgical, Neuropsychological and  Perioperative: Clinical, Surgical | 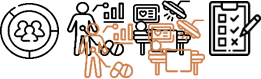 |

**eTable 7.** Performance difference in the area under the receiver operating characteristic curve between models with additional intervention allocation information: 1000 permutation tests.

| *P* | 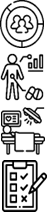 | 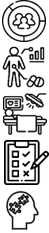 | 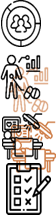 | 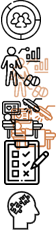 |
| --- | --- | --- | --- | --- |
| 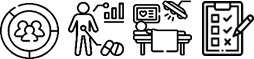 | 1.0000 | 0.7522 | 0.1928 | 0.2158 |
| 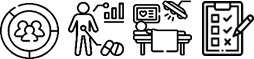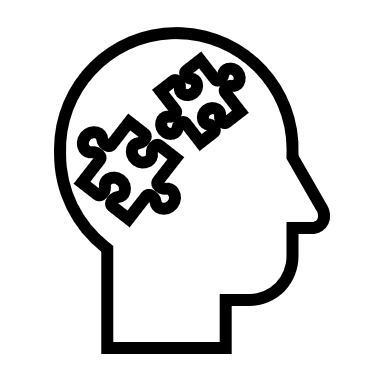 | 0.7522 | 1.0000 | 0.2977 | 0.3626 |
| 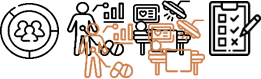 | 0.1928 | 0.2977 | 1.0000 | 0.9161 |
| 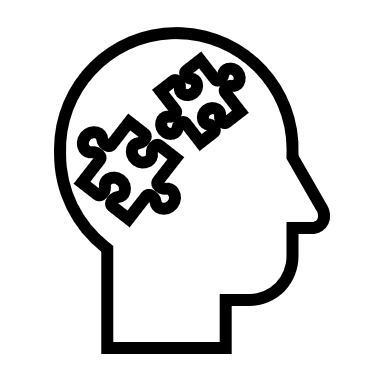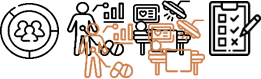 | 0.2158 | 0.3626 | 0.9161 | 1.0000 |

List of icons:

| Preoperative: Demographic, Clinical, Surgical, Neuropsychological | 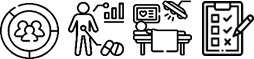 |
| --- | --- |
| Preoperative: Demographic, Clinical, Surgical, Neuropsychological, Intervention allocation | 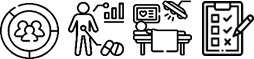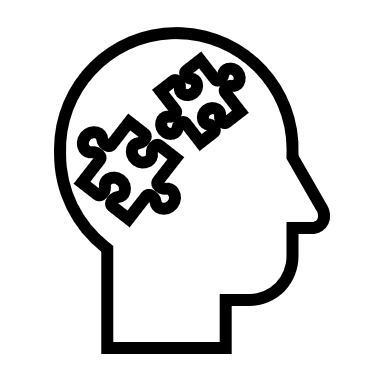 |
| Preoperative: Demographic, Clinical, Surgical, Neuropsychological and  Perioperative: Clinical, Surgical | 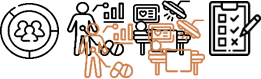 |
| Preoperative: Demographic, Clinical, Surgical, Neuropsychological, Intervention allocation and Perioperative: Clinical, Surgical | 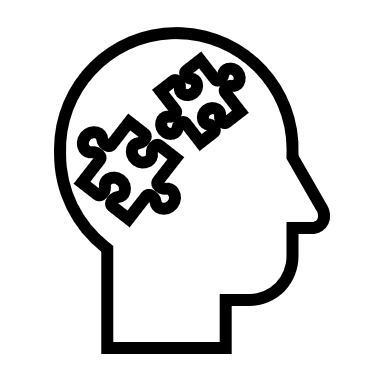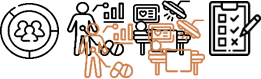 |
